# Longitudinal analysis of B- and T-cell responses to SARS-CoV-2 recombinant S-protein vaccine S-268019-b in phase 1/2 priming and booster study

**DOI:** 10.1101/2024.10.01.24314659

**Authors:** Masaya Fujitani, Xiuyuan Lu, Ryo Shinnakasu, Takeshi Inoue, Yujiro Kidani, Naomi M. Seki, Satoru Ishida, Shungo Mitsuki, Takeshi Ishihara, Miwa Aoki, Akio Suzuki, Koji Takahashi, Masahiro Takayama, Takeshi Ota, Satoshi Iwata, Risa Yokokawa Shibata, Takuhiro Sonoyama, Mari Ariyasu, Ayumi Kitano, Tommy Terooatea, Jordan Kelly Villa, Kazuo Yamashita, Sho Yamasaki, Tomohiro Kurosaki, Shinya Omoto

## Abstract

The durability of vaccine-induced immune memory to severe acute respiratory syndrome coronavirus 2 (SARS-CoV-2) is crucial for preventing infection, especially severe disease. This follow-up report from a phase 1/2 study of S-268019-b (a recombinant spike protein vaccine) after homologous booster vaccination confirms its long-term safety, tolerability, and immunogenicity. Booster vaccination with S-268019-b resulted in an enhancement of serum neutralizing antibody (NAb) titers and a broad range of viral neutralization. Single-cell immune profiling revealed persistent and mature antigen-specific memory B cells and T follicular helper cells, with increased B-cell receptor diversity. The expansion of B- and T-cell repertoires and presence of cross-reactive NAbs targeting conserved epitopes within the receptor-binding domain following a booster accounted for the broad-spectrum neutralizing activity. These findings highlight the potential of S-268019-b to provide broad and robust protection against a range of SARS-CoV-2 variants, addressing a critical challenge in the ongoing fight against coronavirus disease 2019 (COVID-19).

## INTRODUCTION

Severe acute respiratory syndrome coronavirus 2 (SARS-CoV-2) infection and the ensuing coronavirus disease 2019 (COVID-19) have caused 776 million confirmed cases and 7.1 million deaths worldwide, with 33.8 million cases and 74,700 deaths in Japan as of July 2024, with the number of infections continuing to rise.^1^ The timely development of vaccines has been pivotal in controlling the pandemic, with approximately 67% of the world’s population having received at least one dose of a COVID-19 vaccine as of June 2024.^2^ However, vaccine shortages remain an issue in low- and middle-income countries.

The rapid evolution of SARS-CoV-2, leading to the emergence of variants of concern, poses significant challenges for developing vaccines that elicit a broad antibody response. The World Health Organization has identified five such variants: Alpha (B.1.1.7), Beta (B.1.351), Gamma (P.1), Delta (B.1.617.2), and Omicron (B.1.1.529).^3^ Furthermore, vaccine-induced neutralizing antibodies (NAbs) wane over time, reducing the effectiveness of primary vaccinations against SARS-CoV-2 infection.^4–6^ Similarly, immunity following natural infection also wanes over time.^7^ Hence, booster doses have been initiated worldwide, creating a substantial and steady demand for COVID-19 vaccines that are safe and effective against new variants.

The NAb titers serve as an immune correlate of protection over a few months after vaccination.^8,9^ A third booster dose of a vaccine encoding original Wuhan Spike protein induced potent NAb titers against Omicron, providing high protection from infection.^10–13^ Durable immunity is supported by B-cell responses, which include long-lived plasma cells and memory B (Bmem) cells that can rapidly respond to re-exposure.^14^ SARS-CoV-2 infection induces humoral memory responses that confer protection via preexisting antibodies from long-lived plasma cells and recalled antibodies from Bmem cells.^15^ While antibodies against SARS-CoV-2 decline over several months after vaccination and/or infection, Bmem cell responses either rise or are sustained over a period, indicating their crucial role in long-term immunity.^8,16–18^ Current evidence suggests that recall responses are important for protective immunity, especially in modulating disease severity and resolving infection.^19^

T-cell responses crucially contribute to the affinity maturation and longevity of antibodies.^20–22^ Notably, patients with severe COVID-19 exhibited higher antibody titers than those with mild or moderate disease, suggesting a correlation between antibody titers and disease severity.^23–25^ In contrast, SARS-CoV-2–specific CD4^+^ and CD8^+^ T-cell responses were associated with reduced disease severity.^26^ However, the relationship between disease severity, T-cell responses, and antibody titers over time remains unclear, necessitating longitudinal analysis of SARS-CoV-2–specific immune responses at multiple time points to understand the factors that influence T-cell sustainability and the disparity between T-cell and antibody responses.

S-268019-b is a recombinant protein vaccine comprising the S-910823 antigen, a modified recombinant spike protein of SARS-CoV-2 produced in insect cells using the baculovirus expression vector system, with A-910823, a squalene-based adjuvant in an oil-in-water emulsion formulation.^27^ The vaccine received approval from the Ministry of Health, Labor and Welfare in June 2024.^28^ The immunogenicity and safety of this vaccine are proven by a series of clinical trials. A phase 1/2 study (*n* = 60) in Japan demonstrated the safety and immunogenicity of S-268019-b in a primary vaccination series.^29^ Further, T-cell responses showed that S-268019-b induced antigen-specific polyfunctional CD4^+^ T-cell responses, reflected in interferon-gamma (IFN-γ), interleukin 2 (IL-2), and IL-4 production on spike protein stimulation. A randomized phase 2/3 study in Japan (*n* = 204) showed that S-268019-b, when used as a booster dose, elicited robust NAb responses and was non-inferior to BNT162b2, an approved mRNA vaccine.^30^ A phase 3 study in Japan (*n* = 1225) further confirmed its superior immunogenicity compared to ChAdOx1nCoV-19 vaccine.^31^

Building on the earlier report of the phase 1/2 clinical study assessing the safety, tolerability, and immunogenicity of a two-dose primary vaccination with S-268019-b in healthy Japanese adults,^29^ we analyzed the humoral and cellular responses induced by S-268019-b vaccine longitudinally. We also investigated longitudinal immune responses and the evolution of B- and T-cell responses to S-268019-b by employing single-cell transcriptomic and epitope sequencing, and B-cell receptor (BCR) and T-cell receptor (TCR) repertoire profiling on peripheral blood mononuclear cells (PBMCs). This comprehensive analysis provides insights into the immunogenicity and safety profile of S-268019-b over a 1-year follow-up period.

## RESULTS

### Participant disposition

The study design and participant flow are illustrated in **Figure 1**. A total of 133 participants were enrolled in the study. Of the 60 randomized participants (24 in the S-268019-b 5 μg group, 24 in the S-268019-b 10 μg group, and 12 in the placebo group; in the same order hereinafter), 44 completed the follow-up up to Day 386 (87.5% [21/24], 91.7% [22/24], and 8.3% [1/12], respectively). Sixteen participants discontinued from the study (12.5% [3/24], 8.3% [2/24], and 91.7% [11/12], respectively), and the reason for the study discontinuation was “withdrawal by subject or the subject’s legally acceptable representative” for all of them.

**Figure 1.**
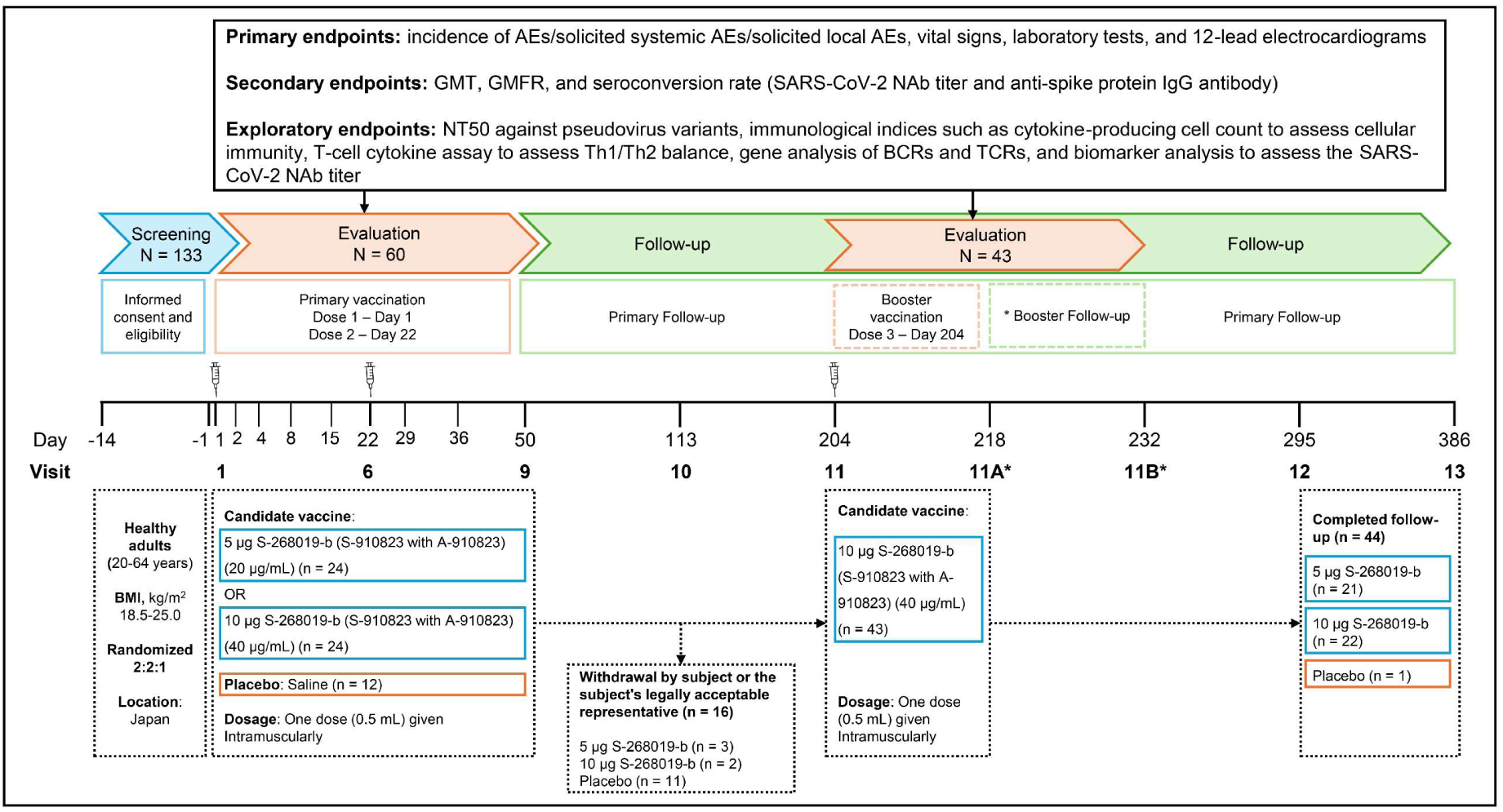
Study design, participant flow, vaccine regimen, and key assessments. Participants were randomly assigned to three study groups and received 0.5 mL of the assigned intervention (5 μg S-268019-b, 10 μg S-268019-b, or placebo) on Days 1 and 22. Follow-up visits during the evaluation period were scheduled on Days 2, 4, 8, 15, 29, 36, and 50. Additional follow-up visits were conducted on Days 113, 204, 295, and 386 during the extended follow-up period. Participants in the S-268019-b 5 μg and 10 μg groups who opted for a booster received a dose of 10 μg S-268019-b intramuscularly on Day 204 and were further followed up on Days 218 and 232. The top section delineates the key assessments conducted throughout the study, including baseline assessments, post-vaccination follow-ups, and final evaluations. Abbreviations: AE, adverse event; BCR, B cell receptor; BMI, body mass index; GMT, geometric mean titer; GMFR, geometric mean fold rise; IgG, immunoglobulin G; NAb, neutralizing antibody; NT50, 50% neutralization titer; SARS-CoV-2, severe acute respiratory syndrome coronavirus 2; TCR, T cell receptor; Th, T helper cell

### Demographic and baseline characteristics

Demographic and baseline characteristics of participants (detailed published previously^29^) are as follows: Nearly half of the participants were male (45.8% in the S-268019-b 5 μg group, 54.2% in the S-268019-b 10 μg group, and 41.7% in the placebo group; in the same order hereinafter). The mean ± standard deviation (SD) age was 43.0 ± 9.9 years, 47.1 ± 9.7 years, and 48.0 ± 8.6 years, respectively. No substantial differences were observed among intervention groups except for habits of alcohol consumption (54.2%, 58.3%, and 16.7%, respectively).

### Safety

The incidences of solicited systemic adverse events (AEs) and solicited local AEs are presented in **Table 1**. No deaths, nonfatal serious AEs (SAEs), AEs leading to discontinuation of study intervention, or AEs of special interest were reported. Except for two solicited systemic AEs (headache and fatigue), all other solicited systemic AEs and solicited local AEs reported in the S-268019-b groups were considered related to the study intervention. Most solicited adverse events were mild to moderate in severity and were transient. Most of the solicited systemic AEs (fever, nausea/vomiting, diarrhea, headache, fatigue, myalgia) occurred within 3 days after the first, second, or booster administration, while most of the solicited local AEs (pain, erythema/redness, induration, swelling) occurred within 2 days after the first, second, or booster administration. Most of the solicited systemic and local AEs resolved within 4 and 6 days, respectively, after each study intervention. For all solicited and unsolicited AEs, the reported outcomes were “recovered/resolved” or “recovering/resolving” for all AEs except for one event each of vertigo and hypertension reported in one participant of the S-268019-b 5 μg group and one event of contusion reported in one participant of the placebo group.

**Table 1.** Incidence of adverse events, treatment-related adverse events, solicited and unsolicited adverse events, treatment-related solicited and unsolicited adverse events in the safety analysis population.

| System Organ Class<sup>a</sup><br>Preferred Term | S-268019-b 5 µg |  |  | S-268019-b 10 µg |  |  | Placebo |  |
| --- | --- | --- | --- | --- | --- | --- | --- | --- |
|  | N = 24 |  |  | N = 24 |  |  | N = 12 |  |
| Number of injections | 1 | 2 | 3 | 1 | 2 | 3 | 1 | 2 |
|  | N = 24<sup>b</sup> | N = 24<sup>b</sup> | N = 21<sup>b</sup> | N = 24<sup>b</sup> | N = 24<sup>b</sup> | N = 22<sup>b</sup> | N = 12<sup>c</sup> | N = 12<sup>c</sup> |
| **Participants with any solicited systemic adverse events** | 10 (41.7) | 21 (87.5) | 19 (90.5) | 13 (54.2) | 19 (79.2) | 20 (90.9) | 2 (16.7) | 2 (16.7) |
| Fever | 0 | 5 (20.8) | 9 (42.9) | 0 | 12 (50.0) | 13 (59.1) | 0 | 0 |
| Nausea/Vomiting | 0 | 8 (33.3) | 8 (38.1) | 4 (16.7) | 11 (45.8) | 11 (50.0) | 0 | 0 |
| Diarrhea | 1 (4.2) | 1 (4.2) | 2 (9.5) | 0 | 0 | 2 (9.1) | 0 | 0 |
| Headache | 6 (25.0) | 13 (54.2) | 16 (76.2) | 7 (29.2) | 15 (62.5) | 17 (77.3) | 0 | 1 (8.3) |
| Fatigue | 7 (29.2) | 21 (87.5) | 18 (85.7) | 7 (29.2) | 18 (75.0) | 18 (81.8) | 2 (16.7) | 1 (8.3) |
| Myalgia | 1 (4.2) | 7 (29.2) | 9 (42.9) | 7 (29.2) | 11 (45.8) | 8 (36.4) | 0 | 1 (8.3) |
| **Participants with any solicited local adverse events** | 22 (91.7) | 23 (95.8) | 20 (95.2) | 21 (87.5) | 22 (91.7) | 21 (95.5) | 3 (25.0) | 3 (25.0) |
| Pain | 21 (87.5) | 23 (95.8) | 18 (85.7) | 21 (87.5) | 22 (91.7) | 21 (95.5) | 3 (25.0) | 3 (25.0) |
| Erythema/Redness | 2 (8.3) | 6 (25.0) | 8 (38.1) | 0 | 6 (25.0) | 5 (22.7) | 0 | 0 |
| Induration | 3 (12.5) | 9 (37.5) | 10 (47.6) | 1 (4.2) | 8 (33.3) | 8 (36.4) | 0 | 0 |
| Swelling | 2 (8.3) | 9 (37.5) | 12 (57.1) | 0 | 5 (20.8) | 5 (22.7) | 0 | 0 |

<sup>a</sup>MedDRA version 24.0.
<sup>b</sup>N is defined as the number of participants who have received first, second, or third administration of study intervention. Number of participants is used as denominator for calculating incidence.
<sup>c</sup>N is defined as the numbers of participants who have received first or second administration of study intervention. Number of participants is used as denominator for calculating incidence.
Data are presented as n/N (%).
Participants with multiple AEs were counted only once within a system organ class and preferred term.

### Immunogenicity achieved with S-268019-b vaccination

#### Anti-spike protein immunoglobulin G (IgG) antibody

After two injections for primary vaccination, the geometric mean titer (GMT) of anti-spike protein IgG antibody increased in the S-268019-b groups (**Figure 2A, Supplementary Figure S1A, B)**. The maximum GMTs after the second dose were observed on Day 36, with highest GMTs in the 10 μg group. After the booster injection of S-268019-b 10 μg on Day 204, GMTs of anti-spike protein IgG antibody increased sharply in both 5 μg and 10 μg groups: on Day 218, GMTs peaked in both the 5 μg and 10 μg groups (**Figure 2A, Supplementary Figure S1A)**. GMTs decreased over time but remained elevated on Day 386 in both groups.

**Figure 2.**
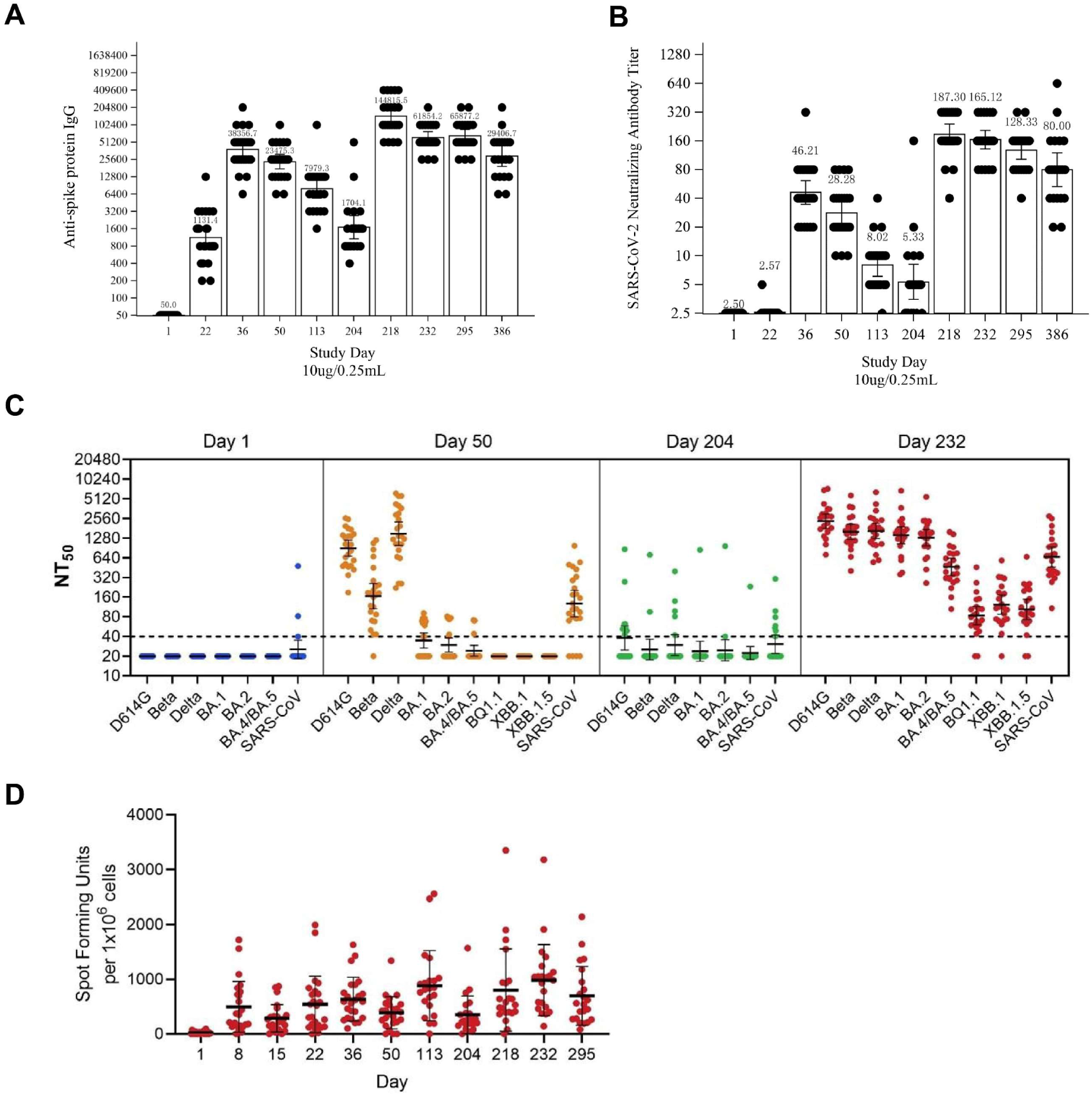
Immunogenicity of S-268019-b (10μg group). See also Figure S1. **(A)** and **(B)** GMTs of anti-spike protein IgG antibodies and SARS-CoV-2 neutralizing antibodies, respectively, in participants receiving a 10 μg dose of S-268019-b. Each bar represents the GMT (error bars indicate a 95% CI). Each point represents the antibody titer in an individual. **(C)** Serum neutralizing antibody titers against various SARS-CoV-2 subvariants. Each horizontal line represents the GMT (error bars indicate a 95% CI). Each circle represents the neutralizing antibody titer in an individual. The dotted line indicates the lower limit of detection. **(D)** ELISPOT analysis showing the number of IFN-γ–producing cells after vaccination. Group = Actual intervention in the primary series. Group names show the antigen content (μg)/adjuvant content (mL). Results are based on samples collected at specified time points post-vaccination. Abbreviations: ELISPOT, enzyme-linked immunospot; GMT, geometric mean titer; NT50, 50% neutralization titer; SARS-CoV-2, severe acute respiratory syndrome coronavirus 2.

After two primary vaccination injections, geometric mean fold rise (GMFR) (95% confidence intervals [CIs]) of anti-spike protein IgG antibody titer on Day 36 were 527.00 (375.27–740.10) in the 5 μg group, 767.13 (555.95–1058.53) in the 10 μg group, and 1.00 (–) in the placebo group (**Supplementary Table S1)**. On Day 50, GMFRs were 287.35 (206.47– 399.91), 469.51 (351.21–627.65), and 1.33 (0.90–1.98), respectively. After a booster on Day 204, GMFRs on Day 218 were 3565.78 (1937.30–6563.14) in the 5 μg group and 2896.31 (2092.45–4008.99) in the 10 μg group. By Day 386, GMFRs decreased in both groups.

#### SARS-CoV-2 neutralizing antibody

Similarly, the GMT of SARS-CoV-2 NAb increased substantially in both the S-268019-b groups after two injections for primary vaccination (**Figure 2B, Supplementary Figure S1C, D)**. The maximum GMTs after the second dose were observed on Day 36, with the highest GMT in the 10 μg group. After a booster injection of S-268019-b 10 μg on Day 204, GMTs of SARS-CoV-2 NAb increased in both the 5 μg and 10 μg groups (**Figure 2B, Supplementary Figure S1C)**. GMTs decreased over time but remained higher than baseline on Day 386.

The GMFR (95% CIs) of SARS-CoV-2 NAb titer were 14.67 (10.25–21.01) in the 5 μg group, 18.49 (13.89–24.61) in the 10 μg group, and 1.00 (–) in the placebo group on Day 36 (**Supplementary Table S1)**. On Day 50, GMFRs were 8.48 (6.14–11.70), 11.31 (8.61– 14.86), and 1.00 (–), respectively. After a booster injection on Day 204, GMFRs in the 5 μg and 10 μg groups were 71.01 (51.54–97.85) and 74.92 (58.36–96.17), respectively, on Day 218. By Day 386, GMFRs were 23.52 (14.20–38.95) and 32.00 (21.29–48.10), respectively.

#### Neutralization against SARS-CoV-2 subvariants

We next evaluated NAb in sera at Days 1, 50, 204, and 232 against pseudoviruses bearing S protein from ancestral D614G, Beta, and Delta subvariants (**Figure 2C)**. The NAb titer against Omicron subvariants declined compared to that against D614G, Beta, and Delta prior to booster, but markedly increased after boosting with S-268019-b as observed at Day 232.

#### IFN-γ–producing cells

After the first injection (primary vaccination), the number of spots/1.0 × 10^6^ PBMCs increased substantially from Day 1 to Days 8, 15, and 22 in the S-268019-b groups (**Figure 2D**). After the second injection, the number of spots/1.0 × 10^6^ cells rose further at Days 36, 50, and 113. A similar upward trend was seen after the booster vaccination on Day 204. The placebo group consistently had fewer cytokine-producing cells than the S-268019-b groups (data not shown).

#### T cells producing T-helper cell 1 (Th1)/Th2 cells

After primary vaccination, the mean proportion of T cells producing Th1 cells (IFN-γ and IL-2) significantly increased from Day 1 to Day 36 in both S-268019-b groups (**Supplementary Figure S1E)**. Similarly, the mean percentage of T cells producing Th2 cells (IL-4 and IL-5) also increased from Day 1 to Day 36. After the booster vaccination, the percentages of T cells producing Th1 and Th2 cells increased from Day 204 to Day 218 in both dosage groups. The placebo group consistently had fewer T cells producing Th1 and Th2 cells at all time points (data not shown).

### Single-cell transcriptome analysis of B cells and BCR repertoire analysis

Immunogenicity evaluation revealed the increase and persistence of NAb titers, as well as an expansion of their breadth, following booster vaccination. These results suggested the contribution of Bmem cell recall and B-cell maturation. To further investigate this, we performed single-cell transcriptome analysis of B cells and BCR repertoire analysis using linking B-cell receptor to antigen specificity through sequencing (LIBRA-seq)^32^ (**Figure 3A)**.

**Figure 3.**
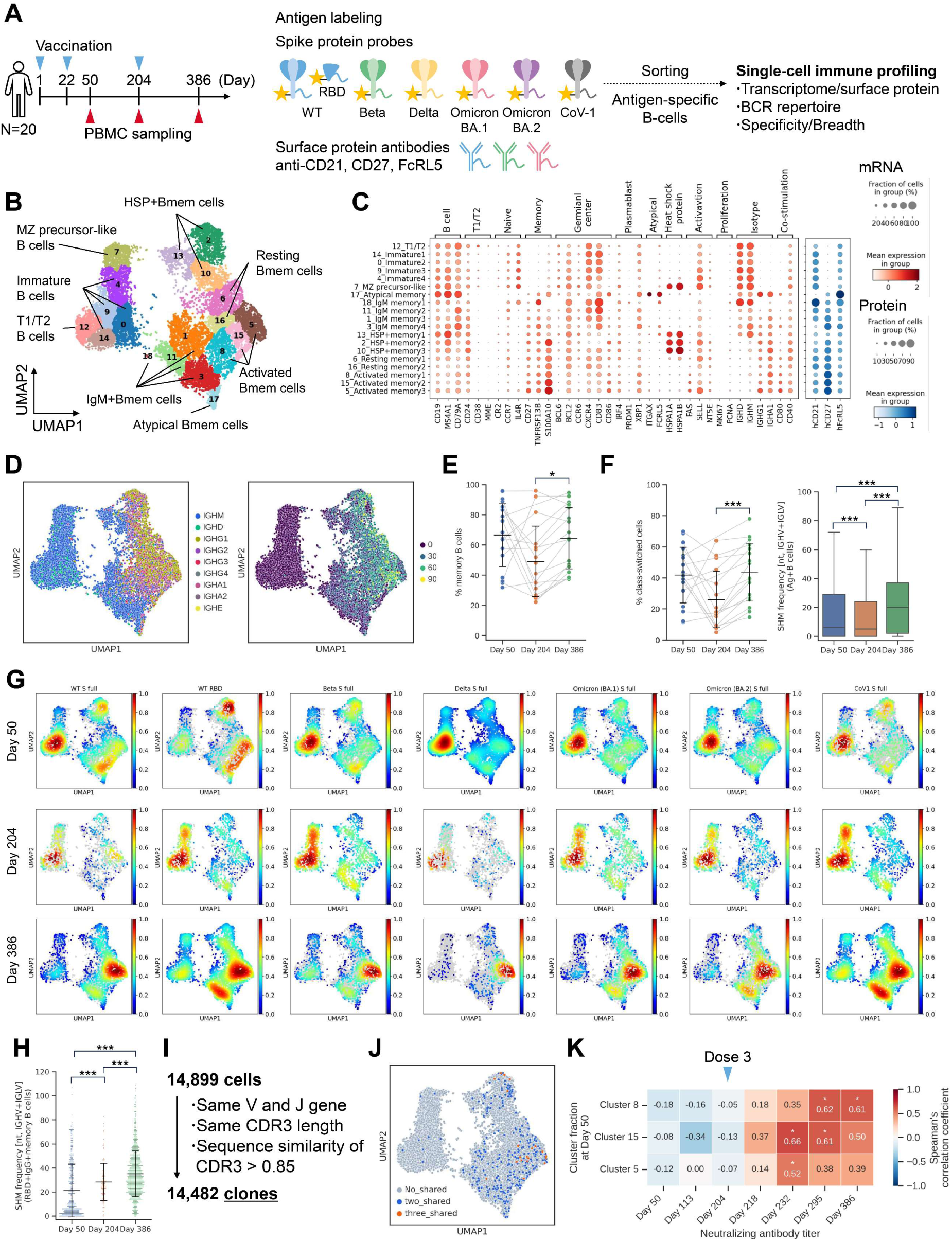
Single-cell analysis of B cells. See also Figure S2 and S3. **(A)** Experimental design for the single-cell analysis. **(B)** UMAP of antigen-specific B cells based on gene expression profiles. **(C)** Bubble plots representing marker gene expression levels, showing mRNA expression (left) and surface protein expression (right) across identified clusters. The size of the bubble represents a fraction of cells expressing the transcript, whereas color is indicative of relative expression. **(D)** Heavy chain isotype (left) and SHM number of IGHV and IGLV (right) visualized on UMAP. **(E)** Fraction of Bmem cells. Each point represents the value for each donor. The connected lines indicate a sample derived from the same donor. Each bar represents mean ± SD. **(F)** Distribution of heavy chain isotype and SHM number across time points. (Left) Fraction of class-switched cells (IgG, IgA, and IgE) per donor. Each point represents the value for each donor. The connected lines indicate samples derived from the same donor. Each bar represents mean ± SD. (Right) Box plot of SHMs in antigen-specific B cells. **(G)** Cell density map illustrating the distribution of probe binding B cells across each time point. The color bars indicate cell density. **(H)** The number of SHMs in RBD-specific IgG Bmem cells. Each point represents the value for each cell. Each bar represents mean ± SD. **(I)** Definition of B-cell clones. **(J)** Clones shared across multiple time points during the study period. No_shared (gray): Clones detected at only one time point, Two_shared (blue): Clones detected at two time points, Three_shared (orange): Clones detected at three time points. **(K)** Correlation analysis. The numbers inside the boxes represent the Spearman’s correlation coefficients between subtype ratio at Day 50 and neutralizing antibody titers. Statistical analyses were performed using Spearman’s rank-order correlation test for (G), pairwise Wilcoxon signed rank test with multiple comparison correction for (E) and (F left), and Tukey’s multiple comparison test for (F right) and (H) (\**p*<0.05, \*\**p*<0.005, \*\*\**p*<0.0005). Abbreviations: Bmem, memory B cells; IgG, immunoglobulin G; RBD, receptor-binding domain; SD, standard deviation; SHM, somatic hypermutation; UMAP, uniform manifold approximation and projection.

LIBRA-seq was conducted using six spike protein probes and wild type (WT) receptor-binding domain (RBD) probes (**Supplementary Figure S2A)**. PBMC samples collected from 20 donors at Day 50, 204, and 386 were labeled with antigen probes. Antigen-specific B cells were sorted based on allophycocyanin (APC)^+^PE^+^ fraction and subjected to sequencing (**Supplementary Figure S2B)**. Negative controls were treated similarly with APC^−^PE^−^ fraction. Transcriptome analysis identified multiple B-cell clusters (**Figure 3B)**. Bmem cells were mapped to the right cluster, which contained subtypes such as activated Bmem cells, resting Bmem cells, IgM Bmem cells, heat shock protein (HSP)^+^ Bmem cells, and atypical Bmem cells (**Figure 3C)**. Activated Bmem cells and resting Bmem cells exhibit class switch and accumulation of somatic hypermutations (SHMs), indicating their status as mature Bmem cells (**Figure 3D)**. The left cluster consisted of immature B cells, which contained subtypes such as T1/T2 stage B cells and marginal zone precursor-like cells (**Figure 3B)**.

To evaluate the B-cell response over time following priming and booster vaccination, we examined the B-cell profiles at each time point. Bmem cells were induced at Day 50 and accounted for more than half of the total population (**Figure 3E, Supplementary Figure S3A)**. RNA-velocity analysis also showed the flow towards the Bmem cell cluster at Day 50 (**Supplementary Figure S3B)**. At Day 204, six months after the priming vaccination, there was a trend of decreased proportion of Bmem cells, but they were still detectable. At Day 386, six months after the booster vaccination, the proportion of Bmem cells was higher than that at Day 204. These findings demonstrate an increase in long-lived Bmem cells following booster vaccination. From the perspective of B-cell maturation, the number of SHMs decreased at Day 204, accompanied by an increase in the population of immature B cells.

However, on Day 386, an increase in the proportion of class-switched cells and the number of SHMs was seen (**Figure 3F**, **Supplementary Figure S3C)**. This result indicates that booster vaccination induced germinal center (GC) reactions and further B-cell maturation. In terms of BCR diversity, although not statistically significant, the Shannon index was numerically higher on Day 386 compared to Day 204, suggesting the appearance of novel clonotypes due to the booster vaccination (**Supplementary Figure S3D)**. However, the usage pattern of V genes was similar across time points (**Supplementary Figure S3E**).

Next, to determine which strains the induced antigen-specific B cells bind to, we analyzed the cell density for each antigen probe. As a result, WT RBD-specific Bmem cells were induced on Day 50 (**Figure 3G)**. Furthermore, the Bmem cells showed an increased number of SHMs at Day 204, indicating that the cells induced by priming vaccination matured further within the six-month period (**Figure 3H)**. Additionally, there was a further increase in the number of SHMs after the booster vaccination. Mutant S full-binding Bmem cells were partially present after priming vaccination but increased for all evaluated antigens at Day 386 (**Figure 3G**).

Moreover, tracking clones based on BCR sequences revealed the presence of Bmem cell clones that were detected at all three time points, indicating the existence of Bmem cell clones maintained for one year (**Figure 3I, J)**. Since the presence of RBD-specific Bmem cells after priming vaccination and the identification of long-lived clones suggest the potential for recall of Bmem cells after booster vaccination, we examined the relationship between the B-cell population after priming vaccination and serum NAb titers (**Figure 3K, Supplementary Figure S3F)**. A positive correlation was observed between the proportion of activated Bmem cell clusters 5, 8, and 15 at Day 50 and NAb titers after the booster vaccination (Day 218, 232, 296, and 386), suggesting the potential contribution of Bmem cells induced by priming vaccination to the NAb titers after the booster vaccination.

B-cell single-cell analysis demonstrated the induction of antigen-specific Bmem cells by S-268019-b priming vaccination. Furthermore, the booster vaccination promoted B-cell maturation and increase of long-lived Bmem cells with antigen specificity including the Omicron variant.

### Diverse epitopes and enhanced neutralization by induced Bmem cells

Single-cell analysis showed the induction of antigen-specific Bmem cells and B-cell maturation. Next, we investigated the function of the identified Bmem cell BCRs. Clusters 5 and 15 contained RBD-specific Bmem cells at Day 50. Moreover, these clusters showed a positive correlation between their proportion at Day 50 and the serum NAb titer after the booster vaccination, suggesting a potential contribution to the neutralizing activity post-booster vaccination. Additionally, these clusters exhibited a high presence of shared clones at multiple time points. Based on this, we generated monoclonal antibodies (mAbs) from selected clones within clusters 5 and 15, and their antibody titers, neutralizing activities, and epitope identification were determined (**Figure 4A)**. Enzyme-linked immunosorbent assay (ELISA) and pseudovirus neutralization assays revealed activity against WT/D614G, BA.1, XBB.1.5, and CoV1, as well as the presence of cross-reactive antibodies (**Figure 4B, Supplementary Table S2)**. Interestingly, the proportion of cross-reactive antibodies among antigen-reactive antibodies was higher at Day 204 (17/21 = 81.0%) and Day 386 (15/17 = 88.2%) compared to Day 50 (12/27 = 44.4%) (**Figure 4C [left])**. Similarly, the proportion of cross-reactive NAbs was higher at Day 204 (9/17 = 52.9%) and Day 386 (8/12 = 66.7%) compared to Day 50 (1/11 = 9.1%) (**Figure 4C [right])**. Moreover, analysis of time point-shared 9 clones revealed an increase in SHM and trends of increased activity and breadth (**Supplementary Figure S4A**).

**Figure 4.**
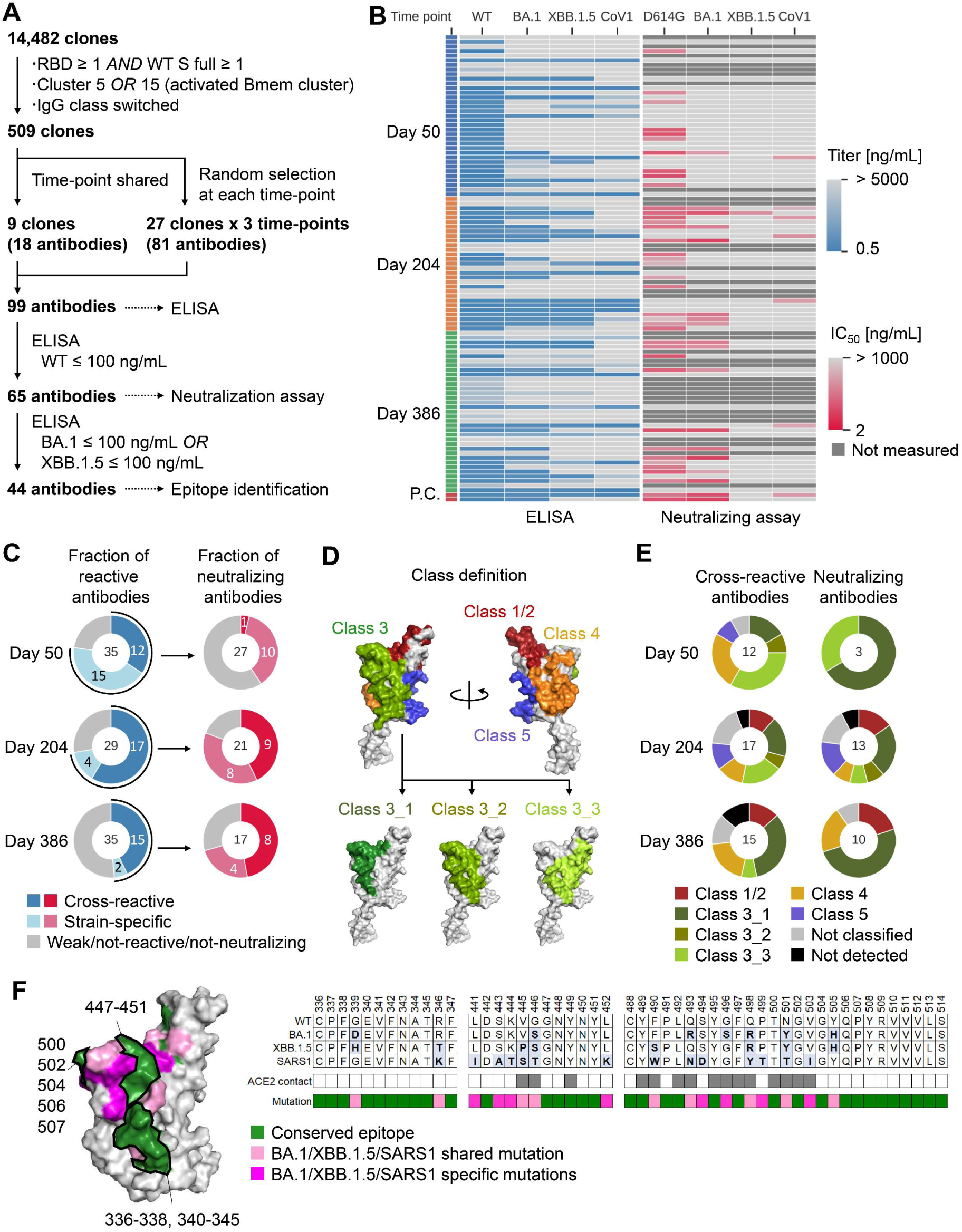
Evaluation of mAbs derived from Bmem cell BCRs. See also Figure S4. **(A)** Flowchart illustrating the selection process of monoclonal antibodies. **(B)** Heat map depicting ELISA results and pseudo-results for 99 antibodies and 2 positive control (P.C.) antibodies (S2K146 and LY-CoV1404). Dark gray cells in the heatmap of neutralizing assay represent unmeasured antibodies. **(C)** Summary pie chart presenting ELISA results and pseudovirus assay results. The numbers within the circles represent the number of antibodies. (Left) The distribution of antigen-binding antibodies is determined based on ELISA results. Antibodies were categorized as follows based on the number of strains with titers of 100 ng/mL or below: cross-reactive: two or more strains, strain-specific: only one strain, weak/not-binding: none of the strains. (Right) The distribution of neutralizing antibodies determined based on pseudovirus assay results. The IC50 threshold for categorization is 1000 ng/mL. **(D)** Definition and classification of RBD epitope classes. **(E)** Classification results of identified epitopes, categorizing them based on their interaction profiles with monoclonal antibodies. The numbers within the circles represent the number of antibodies. **(F)** Epitope mapping of Ab31, a representative antibody of class 3_1. (Left) mapping result to WT RBD structure. The colored residues represent the conserved residues (green), variant shared mutations (magenta), and variant specific mutation (pink) respectively. (Right) Amino acid sequences of Ab31 epitope. Grey cells in ACE2 contact row indicated ACE2 contact amino acid. Abbreviations: ELISA, enzyme-linked immunosorbent assay; IC50, half maximal inhibitory concentration; RBD, receptor-binding domain; WT, wild type.

The epitopes of the 44 antibodies that showed cross-reactivity in ELISA were identified using hydrogen deuterium exchange (HDX)-MS, and epitope classification was performed. The epitope classes were defined as class 1/2, 3, 4, and 5, with reference to the epitopes of known antibodies (**Figure 4D, Supplementary Table S3)**.^33–38^ If an epitope did not fit into any of these classes, it was classified as “Not classified”. It should be noted that class 1 and 2 were combined into a single class due to difficulties in distinguishing them using HDX-MS. Since multiple patterns were detected for Class 3, it was further divided into three subclasses. As a result, a diverse range of epitopes was induced from class 1/2 to 5, with a notable abundance of class 3 (**Figure 4E [left], Supplementary Figure S4B)**. Among the NAbs, class 3 proportion was the highest (**Figure 4E [right]**). Among the evaluated antibodies, class 3 was the most prevalent, particularly class 3_1. The antibodies classified as class 3_1 such as Ab31 contained conserved regions within the epitopes (**Figure 4F, Supplementary Figure S4C**).

The evaluation of mAbs confirmed the functionality of the identified Bmem cell BCRs.

Moreover, the proportion of Bmem cells expressing BCRs that exhibit cross-reactive neutralizing activity increased during the six-month period after priming vaccination and booster vaccination, and these BCRs recognized epitopes containing conserved regions.

### Long-lasting T-cell memory and conserved epitopes post-vaccination

T-cells play an important role in B-cell maturation via the GC reaction. Since mature Bmem cells specific to RBD were already present at Day 50, we performed single-cell analysis of T-cells from Day 1 (pre-vaccination) and Day 50 samples. To evaluate the profile of antigen reactive T-cells, PBMCs were collected, labeled with CellTrace violet™, stimulated with S peptide pools that cover S protein of Wuhan strain as well as mutations of Delta and Omicron (BA.1 and BA.2) strains, and sorted for proliferating T-cells for single-cell and epitope analysis (**Figure 5A, Supplementary Figure S5A)**. Transcriptome analysis detected naive T cells, central memory T cells (Tcm), effector memory T cells (Tem), effector T cells (Teff), circulating T follicular helper cells (cTfh), T regulatory cells (Treg), mucosal associated invariant T cells, gamma-delta T cells (gdT), and Th17 cells (**Figure 5B, C)**. The examination of T-cell subtypes at each time point revealed that the population of differentiated T cells such as cTfh cells and memory T (Tm) cells increased after priming vaccination (**Figure 5D, E, Supplementary Figure S5B)**. Interestingly, antigen-reactive T cells were already present prior to vaccination (Day 1) with the majority being naive T cells. The obtained T cells were clonally identified based on TCR sequences, and TCR epitopes for the top five clones from each donor except 6DA100 (from whom no proliferated T-cells were sorted) were determined (**Figure 5F)**. The top 5 clones of the T-cell receptor α (TRA) and T-cell receptor β (TRB) chains were also detected in the bulk TCR repertoire data (**Supplementary Figure S5C)**. These 95 clones consisted mainly of cTfh cells, Teff cells, Tm (Tcm and Tem) cells, and Treg cells (**Supplementary Table S4)**. Epitope identification was successful for 51 of the 95 clones (**Figure 5G, H, Supplementary Table S4)**. The identified TCR epitopes were mapped to the entire spike protein, with particular epitopes mapping to N-terminal domain (NTD) and S2 were found to be highly conserved in the Omicron variant (evaluated up to XBB.1.5, subsequent variants were not evaluated) (**Figure 5G, H, Supplementary Table S4)**. Furthermore, we found 190 public clones which were shared among donors (**Supplementary Figure S5D)**. These public clones included T follicular helper cells (Tfh) cells, Treg cells, Tm cells, and Teff cells (**Supplementary Figure S5E)**. Most of them were shared by two to three individuals (**Supplementary Figure S5F)**. Epitope identification was successful for 9 of the 24 most prevalent or abundant public clones and revealed recognition of RBD and S2 epitopes (**Supplementary Figure S5G)**. Priming vaccination with S-268019-b induced antigen-reactive T cells, including cTfh cells. Several top clones with identified epitopes targeted conserved regions, suggesting their potential to react to variant strains.

**Figure 5.**
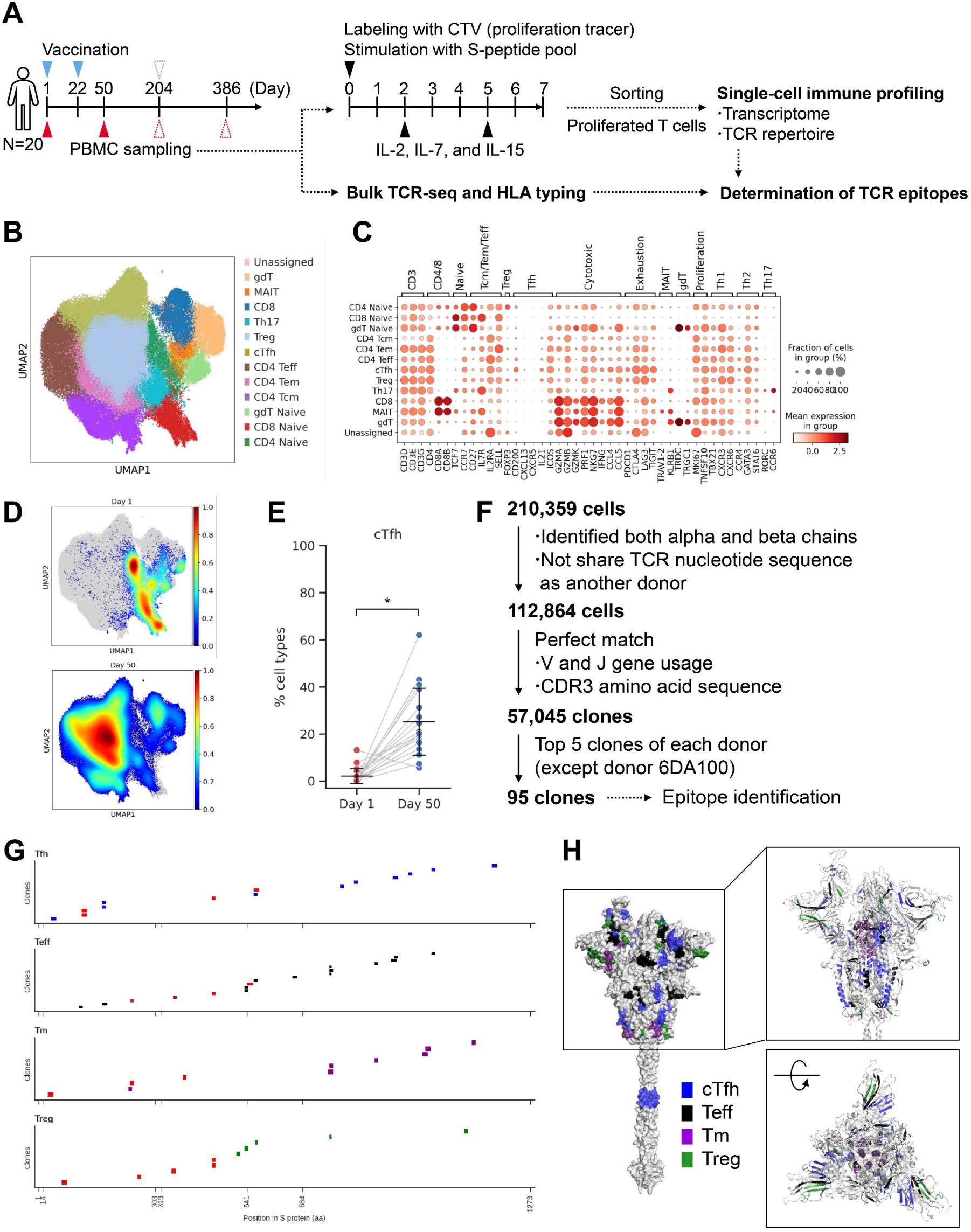
Single-cell analysis of T cells after priming vaccination. See also Figure S5. **(A)** Experimental design for the single-cell analysis. **(B)** UMAP clustering of T cells based on gene expression profiles. **(C)** Bubble plots representing marker gene expression levels. The size of the bubble represents a fraction of cells expressing the transcript, whereas color is indicative of relative expression. **(D)** Density map of T cells at various time points. **(E)** The fraction of cTfh cell. Each point represents the value for each donor. The connected lines indicate samples derived from the same donor. Each bar represents mean ± SD. **(F)** Flow diagram of T-cell clonal definition and selection. **(G)** TCR epitope locations in spike protein. Each short bar indicates the detected epitope region of each clone. The horizontal axis of each graph represents the amino acid residue number of the wild-type spike protein. Red bars indicate the presence of mutations in the Omicron variant (up to XBB.1.5). **(H)** Epitope mapping to the wild-type spike protein structure. The colored residues represent the detected epitopes, and their color indicates T-cell subtypes. (Left) Surface representation. (Upper right) cartoon representation around S1 domain. (Lower right) after 90 degrees rotation. Statistical analyses were performed using pairwise Wilcoxon signed rank test with multiple comparison correction for (E) (\**p*<0.05, \*\**p*<0.005, \*\*\**p*<0.0005). Abbreviations: SD, standard deviation; TCR, T-cell receptor; cTfh, circulating T follicular helper cells; Teff, effector T cells; Tm, memory T; Treg, T regulatory cells; UMAP, uniform manifold approximation and projection.

To confirm the persistence of induced T cells, we performed single-cell analysis on samples collected on Day 204 and Day 386, in addition to the previously collected samples (**Figure 6A)**. By the transcriptome analysis, cTfh cells and Tm cells were detected (**Figure 6B, C, Supplementary Figure S6A)**. The proportion of cTfh cells remained consistent after priming and booster vaccination (**Figure 6D)**. The proportion of CD4-Naïve T cells and CD8-Naïve T cells increased at Day 204 and then decreased at Day 386 (**Supplementary Figure S6B)**. This suggests an increase in the proportion of differentiated T cells after booster vaccination. Since we observed the maintenance of T cells including cTfh cells, we further analyzed the TCR repertoire of the detected 51,121 clones (**Figure 6E)**. The analysis of clonotype proportions between time points revealed that TRBV20-1 was consistently detected at all time points (**Supplementary Figure S6C)**. For TRA, TRAV35-TRAJ42 was predominantly detected after Day 50. The diversity index of clones increased at Day 50 and remained stable until Day 204 and Day 386 (**Figure 6F)**. By subtype, the diversity of effector cells such as cTfh and Tm cells increased at Day 50, while the diversity of naive T cells increased at Day 204 (**Supplementary Figure S6D)**. To assess the persistence of clones, the proportion of the top 16 clones at each time point was aggregated per donor (**Figure 6G)**. As a result, we identified clones that were maintained for six months to one year. Some clones were even maintained from Day 1 before vaccination to after vaccination. We then evaluated the epitope of T cells that persisted for a long period. Among the 51 clones identified in **Figure 5G** at Day 50, 27 clones that were also detected at Day 204 or Day 386 were selected for epitope confirmation. These clones had epitopes in NTD, RBD, and S2 (**Figure 6H**).

**Figure 6.**
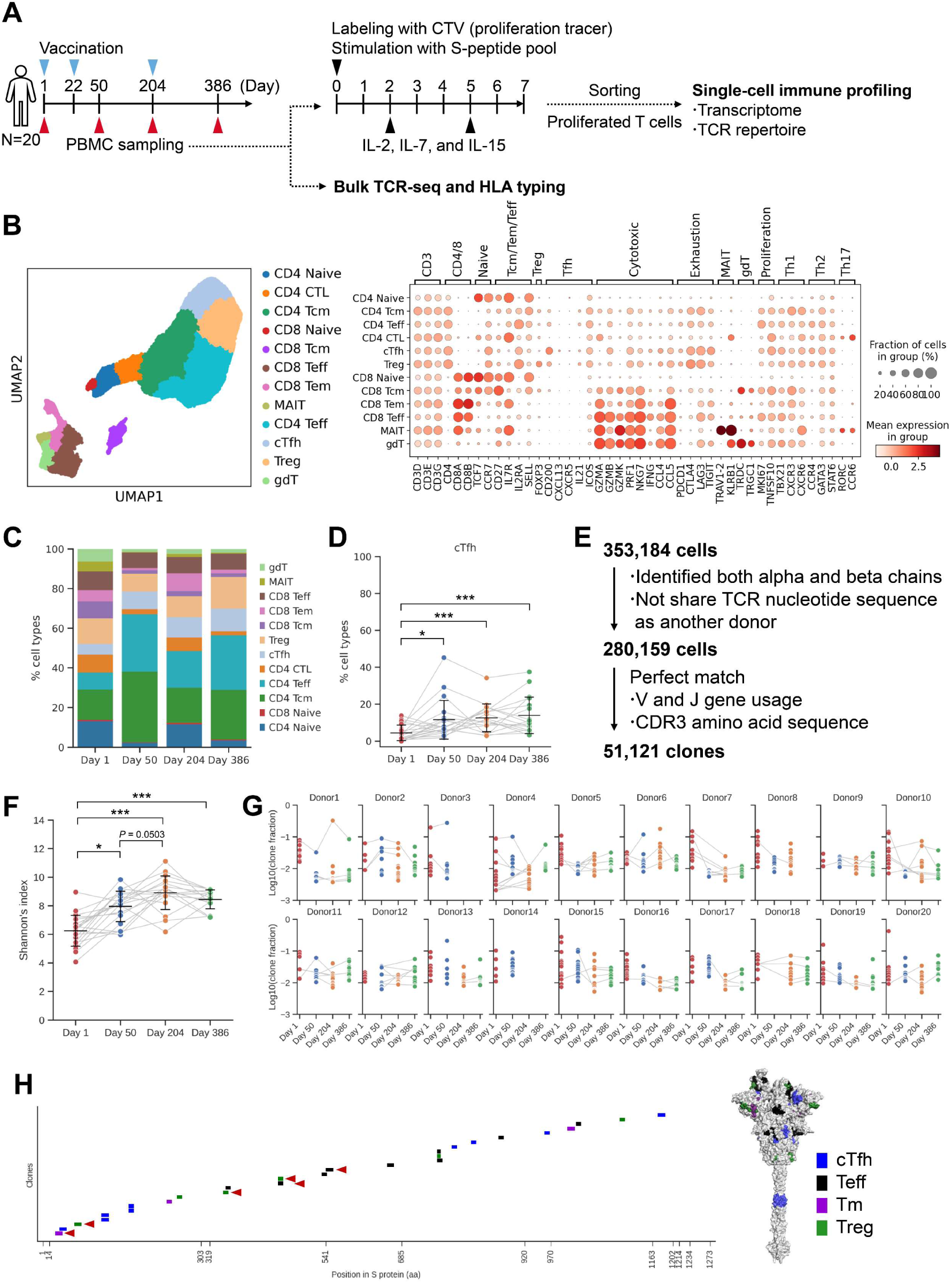
Single-cell analysis of T cells after booster vaccination. See also Figure S6. **(A)** Experimental design for the single-cell analysis. **(B)** UMAP clustering of T cells (left), Bubble plots representing marker gene expression levels (right). The size of the bubble represents a fraction of cells expressing the transcript, whereas color is indicative of relative expression. **(C)** T-cell subtype ratio over time points. The colors indicate subtypes. **(D)** The fraction of cTfh cell. Each point represents the value for each donor. The connected lines indicate samples derived from the same donor. Each bar represents mean ± SD. **(E)** Flow diagram of T-cell clonal definition (left) and the fraction of expanded clones at each time point (right). **(F)** Shannon’s diversity index. Each point represents the value for each donor. The connected lines indicate samples derived from the same donor. Each bar represents mean ± SD. **(G)** The fraction of the top 16 clones at each time point in each donor. Connected lines indicate the same clones. **(H)** Epitope mapping of the persisted clones. (Left) Mapping to sequence. Each short bar indicates the detected epitope region of each clone. The horizontal axis represents the amino acid residue number of the wild-type spike protein. Red arrowheads indicate the presence of mutations in the Omicron variant (up to XBB.1.5). (Right) Mapping to structure. The colored residues represent the detected epitopes, and their color indicates T-cell subtypes. Statistical analyses were performed using pairwise Wilcoxon signed rank test with multiple comparison correction for (D) and (F) (\**p*<0.05, \*\**p*<0.005, \*\*\**p*<0.0005). Abbreviations: SD, standard deviation; cTfh, circulating T follicular helper cells; Teff, effector T cells; Tm, memory T; Treg, T regulatory cells; UMAP, uniform manifold approximation and projection.

Particularly, epitopes of 21 clones present in NTD and S2 showed high conservation. Based on these results, it was observed that T-cell clones, including cTfh cells induced by S-268019-b, were maintained for six months to one year, suggesting their contribution to B-cell maturation through GC reactions and cellular immunity over the long term. The persistent cTfh cells and Tm cells were found to target highly conserved regions as epitopes, indicating their potential to respond to variant strains over an extended period.

## DISCUSSION

In the present phase 1/2 clinical study, we assessed the long-term safety and immunogenicity of primary vaccination with S-268019-b (5 μg or 10 µg) and booster vaccination with S-268019-b (10 μg). S-268019-b was well tolerated and elicited a strong immunogenic response in healthy Japanese adults over a 1-year follow-up. The most common solicited systemic AE across both S-268019-b groups was fatigue, followed by headache and myalgia after the first administration; fatigue, headache, and nausea/vomiting after the second administration; and fatigue, headache, and fever after the booster administration. There were no new safety concerns up to Day 386 (364 days after the second administration or 182 days after the booster administration) in healthy Japanese adults who received three doses of S-268019-b.

The presence of NAbs is currently used as a surrogate indicator of immunity. Preclinical studies on S-268019-b in rodents and monkeys demonstrated enhanced NAbs and cellular immune responses against SARS-CoV-2, including Omicron subvariants.^27,39–42^ Further, S-268019-b was well tolerated as both primary vaccination and booster (regardless of choice of primary vaccination), evident from the low incidence of AEs and treatment-related AEs, aligning with previous studies.^29–31,43–45^ The GMTs of SARS-CoV-2 NAb and anti-spike protein IgG antibodies increased from baseline to Day 50 and remained high on Days 232, 295, and 386, with post-booster levels exceeding those on Day 36. The maximum GMFRs were observed on Day 218, with higher GMFRs of anti-spike protein IgG antibody in the 5 μg group and of SARS-CoV-2 NAb in the 10 μg group. The results indicate that booster vaccination with S-268019-b effectively enhances neutralization against Omicron subvariants and various mutant strains of the virus. A phase 3 study of S-268019-b (10 μg dose, 28 days apart) in Vietnamese participants showed a clear increase in GMTs for SARS-CoV-2 NAbs and anti-spike protein IgG following immunization.^44^ A phase 2/3 study in Japanese adults demonstrated that GMTs for NAbs increased from baseline to Day 15 and Day 29, showing comparable results between S-268019-b and BNT162b2.^30^ An open-label study indicated higher GMTs for NAbs following a booster vaccination with S-268019-b in Japanese adults and elderly participants previously vaccinated with BNT162b2 or mRNA-1273.^45^ In addition to S-268019-b, the development of vaccines against Omicron variants has progressed as part of the COVID-19 vaccine platform. These vaccines are expected to enhance effectiveness against the latest variant strains.

Regarding T-cell responses, the number of IFN-γ–producing cells after the second administration was 10.0–43.2 times higher than before the first dose. After the booster vaccination, IFN-γ–producing cells increased 1.99–3.79 times compared to pre-booster levels and were higher than before the primary vaccination. The data indicated that IFN-γ– producing cells persist for ≥ six months post-primary vaccination. S-268019-b induced a higher percentage of Th1 cells (producing IFN-γ or IL-2) than Th2 cells (producing IL-4 or IL-5) after each vaccination, eliciting a predominantly Th1-mediated immune reaction with minimal Th2 reaction.^29,45^

We conducted single-cell analysis of B and T cells to elucidate the mechanisms of the persistence and broadening of NAb titers. Our results demonstrated the induction and persistence of antigen-specific Bmem cells and cTfh cells following priming vaccination of S-268019-b, as well as B-cell maturation following booster vaccination. Moreover, a positive correlation was observed between activated Bmem cells at Day 50 and the NAb titers after booster vaccination. From these findings, it is suggested that the Bmem cells induced by priming vaccination may contribute to the increase and persistence of NAb titers through recall upon booster vaccination, followed by GC reactions with Tfh cells. Additionally, the recall of Bmem cells possessing conserved epitopes and B-cell maturation may contribute to the increase of cross-reactive clones and the broadening of neutralizing breadth.

A-910823 in S-268019-b is a squalene-based adjuvant. In our previous report, we demonstrated that components such as α-tocopherol can induce Tfh cells and GC B cells in mice.^39^ Evaluations using cynomolgus macaques also confirmed that A-910823 is essential for the booster effect, crucial for inducing NAbs and expanding their breadth.^27^ In this study, we demonstrated the induction of cTfh cells and the increase of class-switch recombination and SHM in B cells after priming and booster vaccination of S-268019-b with A-910823.

This finding suggests that A-910823 strongly induces GC reactions in humans and contributes to persistent humoral immunity. Previous reports indicate the durability of antibody titers in the S-268019-b heterologous booster group compared to individuals who received two doses of mRNA vaccine,^43^ suggesting that persistent humoral immunity may be a characteristic of S-268019-b.

Some immune responses in S-268019-b vaccine recipients resemble those in mRNA vaccine recipients and convalescent individuals. One parameter reflecting B-cell quality is the number of SHMs, essential for high-affinity humoral immune responses. Higher affinity BCRs are selected via Tfh cell help and produced as antibodies by differentiated plasma cells.^46^ Our observations show dynamic changes in B-cell maturation over time. SHMs induced by S-268019-b did not differ substantially from those induced by mRNA vaccines.^47–49^ In terms of the V gene in the BCR, S-268019b recipients showed a higher induction of B cells using specific V genes like IGHV3 23, IGHV3 33, and IGHV4 39. mRNA vaccine recipients and convalescent individuals also exhibited the induction of Bmem cells using these V genes.^50–53^ This suggests that, regardless of vaccine modality, similar V genes may be selected against SARS-CoV-2 to induce antibody production.

From the point of view of TCR epitopes, S-268019-b was found to induce a diverse range of TCR epitopes across the entire S protein. Similarly, mRNA vaccine recipients also exhibit a broad range of induced epitopes, encompassing regions such as NTD, RBD, and S2 regions.^54^ Most identified non-RBD epitopes, including those involving cTfh and Tm, remain conserved even in the Omicron variant, suggesting potential for broad GC responses and cellular immunity against this highly mutated strain, as well as the necessity of including the whole S protein for vaccine antigens.

The epitopes of mAbs were identified using HDX-MS, which allows for high-throughput identification of protein–protein interaction sites at peptide-level resolution.^55^ Combining these epitope data with mutation and antibody assay data offers deeper insights. Class 3_1 shares a similar epitope with LY-CoV1404.^56^ The BA.1+XBB1.5-specific antibody (Ab26,3,80,76,95) epitope region contains four XBB.1.5-specific escape mutations,^57–59^ indicating potential binding sites. Ab31 exhibited neutralizing activity against four strains, with an epitope spanning classes 2, 3, and 4. Residues 336-338, 340-345, 447-451, 500, 502, 504, 506, and 507 were conserved up to CoV1 and included ACE2 binding residues, suggesting this as a potential epitope. The evaluation of mAbs showed an increase in cross-reactive NAbs, like Ab31, targeting conserved regions at Day 204 and Day 386. The induction of such cross-reactive antibodies may contribute to the breadth of NAb titers in serum.

This study provides a comprehensive follow-up of immune responses up to 6 months after booster vaccination with S-268019-b in Japanese adults. This extended follow-up period allows for a thorough assessment of the durability and long-term efficacy of the vaccine, which is crucial for understanding its potential for long-term protection against SARS-CoV-2. The study also uniquely evaluates long-term immune responses at the single-cell level after both priming and booster vaccinations with S-268019-b. This approach provides detailed insights into the behavior and characteristics of individual immune cells, offering a deeper understanding of how S-268019-b influences the immune system over time. A significant novelty of this research lies in the identification of several BCR epitopes that exhibit cross-reactivity and TCR epitopes conserved among various mutant strains. This finding highlights the potential of S-268019-b to provide broad and robust protection against a range of SARS-CoV-2 variants, addressing a critical challenge in the ongoing fight against COVID-19.

The results should be interpreted considering some limitations of the study. Firstly, the small sample size and limited ethnic and racial diversity may limit the generalizability of the findings. Secondly, in the T-cell single-cell analysis, there is a possibility of bias in the cell population due to the expansion culture for 7 days, which was performed to select antigen-responsive T cells. Furthermore, in the evaluation of mAbs, it is important to note that the results represent only a subset of RBD^+^IgG^+^ activated Bmem cells detected. While random selection was performed to avoid bias, these findings were based on small sample size (29–35 antibodies at each time point). Future research should aim to include larger, more diverse populations and explore methods to minimize potential biases in cell culture and analysis processes.

## CONCLUSION

This 1-year follow-up report after homologous booster vaccination in a phase 1/2 study of S-268019-b vaccine confirms the long-term safety, tolerability, and immunogenicity. Booster vaccination of S-268019-b resulted in an enhancement of serum NAb titers and a broad range of viral neutralization. Single-cell analysis revealed the expansion of B- and T-cell repertoire and induction of cross-reactive NAbs targeting conserved epitopes within the RBD following booster vaccination, suggesting a contribution to broad-spectrum neutralizing activity. Overall, the study underscores the long-term efficacy and broad protective potential of S-268019-b against SARS-CoV-2 variants, making it a promising candidate in the ongoing fight against COVID-19.

## Data availability

The human sequence data is being deposited in the JGA (in preparation). The mass spectral raw data is being deposited in the jPOST (in preparation).

## Author contributions

M.F., X.L., R.S., T.I., Y.K., N.M.S., T.S., M.A., K.Y., S.Y., T.K., and S.O. conceived and designed the study. X.L., R.S., T.I., N.M.S., S.M., T.I., M.A., A.S., M.T., T.O., S.I, and R.Y.S. performed experiments and analyzed the data. M.F., A.K., T.T., J.K.V., and K.Y. performed bioinformatics and statistical analysis. M.F., X.L., R.S., Y.K., N.M.S., S.I., S.M., M.T., T.O., R.Y.S., K.Y., S.Y., T.K., and S.O. contributed to the interpretation of the results. T.S., M.A., and S.O. contributed to funding acquisition; supervision, S.Y., T.K., and S.O. supervised the study. M.F., X.L., R.S., Y.K., N.M.S., S.I., S.M., M.A., M.T., T.O., and S.O. drafted the manuscript. All authors contributed to the article and approved the submitted version.

## STAR Methods

### RESOURCE AVAILABILITY

#### Lead contact

Further information and requests for resources and reagents should be directed to and will be fulfilled by the lead contact (Shinya Omoto).

#### Materials availabliltiy

All reagents generated in this study are accessible from the lead contact (Shinya Omoto) upon a reasonable request.

#### Data and code availability

- The data related to the study are reported in the paper and its supplemental information.
- All data reported in this paper will be shared by the lead contact (Shinya Omoto) upon a reasonable request.
- This paper does not report original code.
- Deidentified participant data can be obtained from the lead contact (Shinya Omoto) upon a reasonable request.
- Any additional information required to reanalyze the data reported in this paper is available from the lead contact (Shinya Omoto) upon a reasonable request.

### EXPERIMENTAL MODEL AND STUDY PARTICIPANT DETAILS

This study was a phase 1/2, single-center, randomized, double-blind, placebo-controlled study (jRCT2031210269). The study enrolled healthy Japanese adults with no prior history of vaccination and no documented history of SARS-CoV-2 infection between August 4, 2021, and September 15, 2022. The study included male and female adults aged 20 to 64 years with a body mass index between 18.5 and 25.0 kg/m^2^ at screening. Participants who were unhealthy, likely to be non-compliant, had received any SARS-CoV-2 vaccine (approved or investigational), or had participated in another clinical study within 30 days were excluded.

Those positive for SARS-CoV-2 infection (via SARS-CoV-2 antigen test) or antibodies at screening, or with certain comorbid medical conditions (cardiovascular, respiratory, hepatic, renal, gastrointestinal, endocrine, hematological, or neurological diseases), fever ≥37.5°C on the day of the first administration or having contraindications for intramuscular vaccination or blood collection were also excluded. Other exclusions were hypersensitivity to study interventions or components, or any drug/allergy contraindicating participation (except pollinosis and atopic dermatitis), or ineligibility as determined by the investigator or sub investigator.

### METHODS DETAILS

#### Preparation of vaccines

The S-268019-b vaccine (Shionogi & Co., Ltd., Osaka, Japan) includes the recombinant S-910823 protein, derived from Pango lineage A sequences, and is formulated with the squalene-based adjuvant A-910823. The recombinant S-910823 protein was produced using a baculovirus expression vector system following the previously described method.^27^

#### Vaccine dose and administration

For primary vaccination, S-268019-b (injectable emulsion) was prepared by mixing equal parts of S-910823 (antigen) with A-910823 (adjuvant, oil-in-water emulsion) at the time of use. Specifically, 0.75 mL of A-910823 was taken from the 1 mL vial and combined with 0.75 mL of S-268019 (at 20 μg/mL or 40 μg/mL) to prepare vaccine dose of S-268019-b 5 μg and S-268019-b 10 μg, respectively. Participants received 0.5 mL of their assigned study intervention (S-268019-b 5 μg, S-268019-b 10 μg, or placebo) intramuscularly on Day 1 and Day 22. Those in the S-268019-b 5 μg and 10 μg groups who opted for a booster vaccination (the third study intervention) received a 10 μg dose intramuscularly on Day 204 after unblinding. The study included three groups (S-268019-b 5 μg, S-268019-b 10 μg, and placebo) and the study duration was divided into three periods: Screening (Days –14 to –1), Evaluation (Days 1 to 50), and Follow-up (Days 51 to 386). Participants were randomly assigned to the three study groups, and received 0.5 mL of the assigned intervention (S-268019-b 5 μg, S-268019-b 10 μg, or placebo) on Days 1 and 22. During the evaluation period, follow-up visits were scheduled on Days 2, 4, 8, 15, 29, 36, and 50. Further follow-up visits were scheduled on Days 113, 204, 295, and 386 during the follow-up period.

Participants in the S-268019-b 5 μg and 10 μg groups who opted for a booster received a dose of S-268019-b 10 μg intramuscularly on Day 204 and were followed-up additionally on Days 218 and 232 (**Figure 1)**.

#### Virus neutralization assay

Ancestral WT SARS-CoV-2 virus (WK-521) was isolated and provided by the National Institute of Infectious Diseases, Japan. The cytopathic effect-based virus neutralization assay was performed as previously described.^30^ Briefly, 2-fold serial dilutions of heat-inactivated serum samples were mixed with an equal volume of virus suspension having 100 times the median tissue culture infectious dose, and incubated for 1 h at 37°C. The VeroE6/TMPRSS2 cell suspension was added to the mixture of sample and virus and dispensed into each well of 96-well culture plates. The plates were incubated at 37°C for 5 d with 5% CO2 and then checked for cytopathic effect under a microscope. Virus neutralization titer was defined as the reciprocal of the highest dilution resulting in equal to or more than 50% cell viability.

SARS-CoV-2 pseudovirus generation and neutralization assays were conducted as described previously.^30^ Briefly, lentivirus-based pseudoviruses bearing the S protein were generated in Lenti-X™ 293 T cells (Takara Bio Inc.), and the viral suspensions having the same viral RNA copies/mL were prepared. Two-fold serial dilutions of heat-inactivated sera were mixed with an equal volume of viral suspension and incubated for 1 h at 37°C. The mixtures of sample and virus were added in duplicates to HEK293T stably expressing human ACE2 and TMPRSS2 cells, followed by incubating the plates at 37°C with 5% CO2 for 2 d. Cells were subjected to luciferase assay and the intensity of luminescence was measured by a microplate reader. Percent neutralization was calculated as the difference between relative light units (RLUs) of virus control wells and test sample wells: % Neutralization = 100% × [1 − (mean RLU of duplicate sample wells ÷ mean RLU of virus control wells)]

The dilution factor for achieving 50% neutralization (50% neutralization titer; NT50) was calculated by using the XLfit^®^ software (version 5.3.1.3) (IDBS). When the percentage neutralization was less than 50% at the first dilution, the NT50 was expressed as half of the first dilution factor.

#### Recombinant protein probe preparation

SARS-CoV-2 spike protein sequences were obtained from GenBank (WT: MN994467) and GISAID (Beta: EPI_ISL_768642, Delta: EPI_ISL_1914591, Omicron BA.1: EPI_ISL_6640917, and Omicron BA.2: EPI_ISL_9595859). The spike proteins were introduced with mutations in the furin cleavage site (RRAR to RAAA) with six stabilizing mutations (WT numbering; F817P, A892P, A899P, A942P, K986P, and V987P).

SARS-CoV-1 spike protein sequence was obtained from GenBank (AY310120.1). The spike protein was introduced with two stabilizing mutations (K969P and V970P). The transmembrane domain was excluded in all spike protein. All spike proteins with a T4 foldon trimerization motif, histidine tag, and Avi-tag were codon-optimized for human cells and cloned into the mammalian expression pCMV vector. RBD (amino acids 331-529) with the signal peptide (MIHSVFLLMFLLTPTESYVD), histidine tag, and Avi-tag was cloned into the mammalian expression pCAGGS vector. Recombinant proteins were produced using Expi293F™ cells according to the manufacturer’s protocol (Thermo Fisher Scientific). The spike/RBD expression vector and BirA expression vector were co-transfected with biotin supplement at 100 μM to obtain biotinylated recombinant proteins. The transfected cells were cultured for 5 d. The recombinant proteins were purified using TALON Metal Affinity Resin (Takara Bio Inc.). Fluorescent-labeled protein probes were prepared by conjugating Avi-tag biotinylated spike/RBD proteins and the following fluorochrome-labeled streptavidin at 4:1.5 ratio overnight at 4°C: streptavidin-APC (Thermo Fisher Scientific) and TotalSeq™-C0951 streptavidin-phycoerythrin (BioLegend^®^), or TotalSeq™-C0953 PE streptavidin (BioLegend^®^), or TotalSeq™-C0954 PE streptavidin (BioLegend^®^), or TotalSeq™-C0955 PE streptavidin (BioLegend^®^), or TotalSeq™-C0961 PE streptavidin (BioLegend^®^), or TotalSeq™-C0962 PE streptavidin (BioLegend^®^), or TotalSeq™-C09512 PE streptavidin (BioLegend^®^), or TotalSeq™-C0966 PE streptavidin (BioLegend^®^).

#### 10x Chromium PBMC sample preparation

We prepared 60 samples (20 subjects × 3 time points) for single-cell analysis. The sample preprocessing was conducted in six batches, with 10 samples each. PBMCs were stained with recombinant protein probes. The cells were stained with the following hashtag antibodies: TotalSeq™-C0251 anti-human Hashtag 1 (clone LNH-94, BioLegend^®^), TotalSeq™-C0252 anti-human Hashtag 2 (clone LNH-94, BioLegend^®^), TotalSeq™-C0253 anti-human Hashtag 3 (clone LNH-94, BioLegend^®^), TotalSeq™-C0254 anti-human Hashtag 4 (clone LNH-94, BioLegend^®^), TotalSeq™-C0255 anti-human Hashtag 5 (clone LNH-94, BioLegend^®^), TotalSeq™-C0256 anti-human Hashtag 6 (clone LNH-94, BioLegend^®^), TotalSeq™-C0257 anti-human Hashtag 7 (clone LNH-94, BioLegend^®^), TotalSeq™-C0258 anti-human Hashtag 8 (clone LNH-94, BioLegend^®^), TotalSeq™-C0259 anti-human Hashtag 9 (clone LNH-94, BioLegend^®^), and TotalSeq™-C0260 anti-human Hashtag 10 (clone LNH-94, BioLegend^®^).

CD19^+^B cells were enriched from stained cells using CD19 MACS beads (Miltenyi Biotec) and LS Columns (Miltenyi Biotec). MACS-sorted samples were stained with the following antibodies: BD Horizon™ BV421 Mouse Anti-Human CD19 (clone HIB19, BD Biosciences), BD Pharmingen™ PE-Cy™7 Mouse Anti-Human IgD (clone IA6-2, BD Biosciences), 7-amino-actinomycin D (7-AAD) (BioLegend^®^), TotalSeq™-C0154 anti-human CD27 (clone O323, BioLegend^®^), TotalSeq™-C0181 anti-human CD21 (clone Bu32, BioLegend^®^), TotalSeq™-C0829 anti-human CD307e (FcRL5) (clone 509f6, BioLegend^®^). The stained cells were analyzed with FACSAria™ II flow cytometer (BD Biosciences) and sorted 7-AAD^-^CD19^+^PE^+^APC^+^ fraction as antigen-specific B cells.

#### Single-cell processing and NGS

For single-cell processing and sequence library construction,10x Genomics single-cell immune profiling technology was used. Briefly, the sorted cells were mixed with reagents of Chromium Next GEM Single Cell 5’ Reagent Kits v2 (10x Genomics) and loaded to Chromium Next GEM Chip K Automated Single Cell Kit (10x Genomics) on Chromium Controller (10x Genomics) for Gel bead-in EMulsion (GEM) generation and cDNA synthesis. After cDNA amplification, the library construction was performed using Chromium Single Cell 5’ Library Construction Kit (10x Genomics). VDJ library and Feature barcode library were prepared using Chromium Single Cell Human BCR Amplification Kit (10x Genomics) and 5’ Feature Barcode Kit (10x Genomics), respectively. All procedures were performed according to the manufacturer’s protocol (Chromium Single Cell 5’ Reagent Kits User Guide (v2 Chemistry Dual Index)). Each library was assessed using Qubit Fluorometer (Thermo Fisher Scientific) and Tape Station System (Agilent) to measure the cDNA concentration and fragment size. The prepared libraries were converted to DNBSEQ format libraries using MGIEasy Universal Library Prep Set (MGI) and DNBSEQ-G400RS High-throughput Sequencing Set (MGI). The converted libraries were sequenced using DNBSEQ-G400 (MGI).

#### Single-cell transcriptome & BCR analysis

Raw gene expression matrices and cell surface protein matrices for each run were generated by Cell Ranger pipeline (version 7.1.0, 10x Genomics) with the reference refdata-gex-GRCh38-2020-A provided by 10x Genomics. These count matrices were processed using Python package Scanpy (version 1.8.0) and R package Seurat (version 5.0.0). Briefly, cells with any of the following conditions were excluded from the analysis: <2,000 or >10,000 unique molecular identifier (UMI), <800 or >2,700 detected genes, <20% or >50% of ribosomal protein-derived mRNA, and >3% of mitochondrial genes. Demultiplexing was performed by HashSolo (version 1.3) and VireoSNP (version 0.5.7) with centered log ratio (CLR) normalized cell surface protein matrices. The LIBRA score was calculated by CLR normalization of antigen probe counts. The log normalized gene expression matrices were integrated using Seurat reciprocal principal component analysis method with the first 50 principal components (PCs) obtained from 3,000 highly variable genes. To summarize the integrated dataset, UMAP was conducted using 50 PCs. The Leiden algorithm was used for clustering. B-cell clusters in UMAP were annotated using the following markers: B; CD19^+^, activated Bmem; CR2(CD21)^low^CD27^+^FAS^+^, resting Bmem; CR2(CD21)^+^CD27^+^, HSP^+^ Bmem; HSP^+^CD27^+^, IgM Bmem; IgM^+^CD27^+^, atypical Bmem; ITGAX(CD11c)^+^FCRL5^+^, immature B; IgM^+^CR2(CD21)^+^CD27^-^, T1/T2 stage; CD24^+^CD38^+^MME^+^, marginal zone precursor like; HSP^+^CD24^+^CD38^+^BCL6^+^. RNA velocity analysis was performed using scVelo (version 0.2.5). BCR clonotype assignment was initially performed using Cell Ranger pipeline with the reference refdata-cellranger-vdj-GRCh39-alts-ensembl-5.0.0. Individual BCR contigs were reannotated with IgBLAST (version 1.20.0) based on the IMGT database (downloaded on 4/12/2023). Clones were defined using DefineClones function of Change-O packages of Immcantation pipeline (version 1.3.0). The definition of a clone was as follows: identical V and J gene usage, same complementarity-determining region (CDR)3 length, and the amino acid sequence similarity based on Hamming distance of CDR3 greater than 85%. Clonal diversity analysis was performed using scikit-bio (version 0.5.6) and other analyses and visualization were performed using matplotlib (version 3.5.1), seaborn (version 0.11.2), ggplot2 (version 3.3.6), and dplyr (version 1.0.9). Python and R versions were 3.9.7 and 4.2.0, respectively.

#### mAb selection

We first filtered 509 clones of RBD^+^IgG^+^ activated Bmem cells by the following filters: (1) binding to RBD (RBD LIBRA score >1 and S full trimer WT >1), (2) belonging to cluster 5 or 15, and (3) heavy chain isotype IgG. From these clones, 9 clones (18 BCRs) detected at multiple time points were preferentially selected. From the remaining 500 clones, 27 clones (27 BCRs) at three time points were randomly selected respectively. A total of 99 BCRs were selected.

#### mAb generation

Recombinant mAbs were prepared as previously described.^60^ Briefly, the VH/VL genes of spike-/RBD-binding B-cell clones identified by next-generation sequencing were commercially synthesized and cloned into expression vectors with human IgG1 heavy chain and kappa/lambda light chain.

#### Enzyme-linked immunosorbent assay

The binding activities of mAbs were measured by ELISA. Briefly, 20 μL of 2 μg/mL recombinant each SARS-CoV-2 or SARS-CoV-1 spike RBD protein (AcroBiosystems) in phosphate-buffered saline (PBS) was added to each well of a 384-well assay plate (Thermo Fisher Scientific) and incubated at 4℃ overnight. The next day, the assay plate was washed by PBS-T (PBS with 0.05% v/v Tween-20) and blocked with each well of 80 μL blocking buffer (PBS with 0.05% v/v Tween-20 and 1% w/v bovine serum albumin) for 1.5 h at room temperature. mAbs were diluted in dilution buffer (PBS with 0.05% v/v Tween-20 and 0.1% w/v bovine serum albumin) at a concentration of 5,000 ng/mL at first, and serially diluted at 1:5 ratio for an eight-point dose curve. After removal of blocking buffer, the assay plate was washed by PBS-T and 20 μL diluted mAbs were added to each well and incubated for 1.5 h at room temperature. After the assay plate was washed by PBS-T, 20 μL horseradish-peroxidase–conjugated anti-human IgGFc fragment antibody (Bethyl Laboratories) was added to each well and incubated for 1.5 h at room temperature. After the assay plate was washed by PBS-T, 20 μL 3,3’,5,5’-tetramethylbenzidine (Thermo Fisher Scientific) was added to each well and incubated for 5 min at room temperature. Finally, 20 μL 2N sulfuric acid (H2SO4) was added to each well and the optical density at 450 nm (OD450) was measured using an Envision plate reader (Perkin Elmer). Binding curves were fitted to a 4-parameter logistic model by using TIBCO Spotfire Analyst (version 11.4.3 LTS), and the concentration of mAbs at which OD450 = 0.5 was calculated as ELISA-based titer of mAbs.

#### HDX mass spectrometry

The plasmids for the recombinant SARS-CoV-2 variants were transiently transfected into Expi293F™ cells (Thermo Fisher Scientific) with the ExpiFectamine™ 293 Transfection Kit (Thermo Fisher Scientific) according to the manufacturer’s protocol. After 4–5 d culture in the Expi293™ Expression Medium (Thermo Fisher Scientific), supernatants were collected and passed through a 0.22-μm filter. The recombinant proteins were purified from supernatants by NGL COVID-19 Spike Protein Affinity Resin (Repligen) according to the manufacturer’s protocol. The elute with 100 mM glycine-HCl (pH 2.7) from the resin was immediately neutralized with 1/10 fraction volume of 1 M Tris-HCl pH 8.5 and concentrated using an Amicon Ultra-2 Centrifugal Filter Unit 100 kDa NWMCO (Merck Millipore), in which the elution buffer was exchanged with PBS (GIBCO).

The initiation of deuterium labeling, labeling reaction time, quench reaction time, injection into the UPLC system, and digestion time were controlled fully automatically by the HDX-PAL system (Leap Technologies, Inc.). The chimeric protein used for hydrogen/deuterium exchange epitope mapping included the RBD of SARS-CoV-2 spike protein (amino acids 319-537 of the spike protein described in GenBank: QHD43416.1).

Deuterium buffer was prepared at pH 7.4 with 10 mM PBS in D2O. The RBD protein and each antibody were mixed in a molar ratio of 1:1 and incubated at 37°C for 30 min to form complexes. Unbound RBD samples or complexed samples were diluted 10-fold in either PBS, pH 7.4 (for non-deuterated experiments), or deuterium buffer (for deuterated experiments) and deuterated at 10°C with labeling times of 60, 120, and 240 s. Deuterium-labeled samples were quenched at 0°C for 3 min by adding 1:1 volume of ice-cold quenching buffer (4 mol/L guanidine hydrochloride, 0.2 mol/L glycine hydrochloride, 0.5 mol/L Tris(2-carboxyethyl)phosphine, pH 2.7). The quenched samples were injected into the UltiMate 3000 UPLC system (Thermo Fisher Scientific). Online digestion was performed on an enzyme pepsin column (Waters) at 8°C for 270 s. Digested peptides were captured on a Hypersil Gold trap column (Thermo Fisher Scientific) at 1°C and eluted into Acclaim PepMap300 C18 analytical column (Thermo Fisher Scientific) with 7-minute gradient separation of 10%–35% B (mobile phase A: 0.1% formic acid in water, mobile phase B: 0.1% formic acid in acetonitrile). Mass spectral data were acquired in positive ion mode using an Orbitrap Eclipse™ Tribrid™ Mass Spectrometer (Thermo Fisher Scientific) with the following settings: electrospray voltage +4.0kV, capillary temperature 275°C, resolution for full scan 120000, resolution for MS/MS scan 60000, m/z range 260–2000, scan time 1 s.

Peptide identification was performed with Byos software (version 5.3.44) (Protein Metrics). The HDExaminer (version 3.1.0) (Sierra Analytics) was used to calculate the hydrogen/deuterium exchange ratio of each identified peptide from the MS raw data files of all HDX experiments. All the peptides analyzed with HDExaminer were manually cleaned up to minimize false positive hits. The deuteration difference between unbound/complexed samples (Δ%D) for each peptide was calculated, and peptides meeting the following criteria 1 and 2 were filtered. ^65^ Criterion 1: Δ%D is greater than 5% for two or more adjacent peptides. Criterion 2: The peptide lengths are 4 amino acids or more. The amino acid residues of the SARS-CoV-2 spike protein corresponding to the regions containing the filtered peptides were identified as epitopes of the antibody. All MS data were submitted to the ProteomeXchange Consortium via jPOSTrepo (https://repository.jpostdb.org/) using the dataset identifier (currently in preparation). Based on the epitope information of anti-RBD antibodies,^33–38^ the RBD of SARS-CoV-2 spike protein was classified into class 1/2, class 3, class 4, class 5, and “Not classified”. The identified epitopes of each antibody were classified according to **Supplementary Table S3**.

#### Enzyme-linked immunospot (ELISPOT) assay

IFN-γ ELISPOT analysis was performed ex vivo using human PBMCs as described previously.^31^ Briefly, tests were performed in triplicate and with a positive control (Cell Activation Cocktail, Bio Legend) and a negative control (medium), and the measurement was performed using the measurement kit, Human IFN-γ Single-Color ELISPOT-Rapid (Cellular Technology Limited [CTL]). CTL precoated plate was washed with D-PBS. Per well, 1.0 × 10^6^ cells (for positive control: 5.0 × 10^4^ cells) were stimulated for 19.5–20.5 h with overlapping peptide pools of SARS-CoV-2 S (Miltenyi Biotec). Plates were scanned using ImmunoCapture 7.0.16.1 (CTL) and counted using ImmunoSpot 7.0.30.4 (CTL). Spot counts were displayed as mean values of each triplicate. The spot count values were normalized by the spot count value of negative control by using the following formula: The spot count value = spot count value of peptide pools of SARS-CoV-2 S − spot count value of the negative control.

#### ICS-flow cytometry

Cytokine-producing T cells were identified by intracellular cytokine staining (ICS) as described previously.^31^ Briefly, PBMCs were thawed and rested for 4–5 h in R10 supplemented medium, were restimulated (1.0 × 10^6^ cells per well) with overlapping peptide pools of SARS-CoV-2 S (Miltenyi Biotec) and epitope peptide pools (Shionogi & Co., Ltd.) in the presence of eBioscience™ Protein Transport Inhibitor Cocktail (Thermo Fisher Scientific K.K.) for 16 h at 37 °C. Controls were treated with a dimethyl sulfoxide-containing medium. Cells were stained for viability and surface markers (BV421™ anti-human CD3 [BioLegend^®^]; BV510™ anti-mouse CD4 [BioLegend^®^]; BB515 mouse anti-human CD8 [BD Biosciences]) in staining buffer and Brilliant Stain Buffer Plus (BSB Plus, BD Horizon, according to the manufacturer’s instructions) for 16–20 h in a refrigerator. Next, the samples were fixed and permeabilized using the BD Cytofix/Cytoperm™ Fixation/Permeabilization kit (BD Biosciences), according to the manufacturer’s instructions. IFN-γ (PE-Cy™7 mouse anti-human IFN-γ [BD Biosciences], and BD Horizon™ BB700 Rat Anti-Human interleukin (IL)-2 [BD Biosciences], APC anti-human IL-4 [BioLegend^®^], and PE anti-mouse/human IL-5 [BioLegend^®^]) assessment was performed in the Perm/Wash buffer supplemented with BSB Plus (BD Horizon, according to the manufacturer’s instructions) for 16–20 h in a refrigerator. Samples were acquired on BD FACSCanto™ II (BD Biosciences) and analyzed with the FlowJo software version 7.6.5 (Becton, Dickinson and Company).

#### In vitro stimulation of T cell

In vitro stimulation of T cell was conducted as described previously.^54,61^ Briefly, PBMCs were thawed and washed with RPMI 1640 medium (Sigma) supplemented with 5% human AB serum (GeminiBio), penicillin (Sigma), streptomycin (MP Biomedicals), and 2-mercaptoethanol (Nacalai Tesque). PBMCs were then stained with CellTrace™ Violet Cell Proliferation Kit (ThermoFisher) in accordance with the manufacturer’s instruction. These cells were subsequently stimulated with S peptide pool (1 μg/mL per peptide, JPT) for 7 d, with the addition of human recombinant IL-2 (1 ng/mL, Peprotech), IL-7 (5 ng/mL, BioLegend^®^), and IL-15 (5 ng/mL, Peprotech) supplemented on Day 2 and Day 5. On Day 7, the cells were washed and stained with anti-human CD3 antibodies. The proliferated T cells (CD3^+^CTV^low^) were sorted by cell sorter SH800S (SONY) and used for single-cell analysis.

#### Single-cell transcriptome & TCR analysis

Raw gene expression matrices and cell surface protein matrices for each run were generated by Cell Ranger pipeline (version 7.1.0, 10x Genomics) with the reference refdata-gex-GRCh38-2020-A provided by 10x Genomics. These count matrices were processed using Python package Scanpy (version 1.8.0) and R package Seurat (version 5.0.0). Briefly, cells with any of the following conditions were excluded from the analysis of priming vaccination samples (**Figure 5)**: <100 UMI, <200 detected genes, >40% of ribosomal protein-derived mRNA, and >20% of mitochondrial genes. As the analysis of samples from all time points showed that clusters of cells not expressing TCR were formed due to an increase in the number of cells, the conditions were changed as follows: <1,000 or >5,000 UMI, <1,000 or >2,000 detected genes, <5% or >30% of ribosomal protein-derived mRNA, and >4% of mitochondrial genes. Demultiplexing was performed by HashSolo (version 1.3) and VireoSNP (version 0.5.7) with CLR normalized cell surface protein matrices. The log normalized gene expression matrices were integrated using Seurat reciprocal principal component analysis method with the first 50 PCs obtained from 3,000 highly variable genes. To summarize the integrated dataset, UMAP was conducted using 50 PCs. The Leiden algorithm was used for clustering. T-cell clusters in UMAP were annotated using the following markers: CD4 T; CD3E^+^CD4^+^, CD8 T; CD3E^+^CD8A^+^, naive; TCF7^+^CCR7^+^, Tcm; IL2RA^+^IL7R^+^ CCR7^low^, Tem; IL2RA^−^CCR7^low^, Teff; IL2RA^+^SELL^low^, cTfh; IL21^+^CD200^+^ICOS^+^PDCD1^+^, Treg; FOXP3^+^IL2RA^+^, MAIT; KLRB1^+^TRAV1-2^+^, gdT; TRDC^+^TRGC1^+^, Th17; RORC^+^CCR6^+^,CD4 CTL; CD4^+^GZMA^+^. TCR clonotype assignment was initially performed using Cell Ranger pipeline with the reference refdata-cellranger-vdj-GRCh39-alts-ensembl-5.0.0. Individual TCR contigs were reannotated with IgBLAST (version 1.20.0) based on the IMGT database (downloaded on 4/12/2023). Clones were defined using DefineClones function of Change-O packages of Immcantation pipeline (version 1.3.0). TCR clones were defined by V gene, J gene, and CDR3 amino acid sequence. Clonal diversity analysis was performed using scikit-bio (version 0.5.6) and other analysis and visualization were performed using matplotlib (version 3.5.1), seaborn (version 0.11.2), ggplot2 (version 3.3.6), and dplyr (version 1.0.9). Python and R versions were 3.9.7 and 4.2.0, respectively.

#### Bulk TCR repertoire analysis

RNA extraction was performed using the RNeasy^®^ Mini Kit (Qiagen) according to the manufacturer’s protocol. Centrifugation in the protocol was performed at 10,000 rpm for “≥10,000 rpm” and at 15,000 rpm for “full speed”. Amplification of TCRa and TCRb and library construction were performed using the SMARTer Human TCR a/b Profiling Kit (Takara Bio Inc.) following the manufacturer’s protocol. Purification of amplified libraries was performed using SPRIselect (Beckman Coulter). The quality of extracted RNA and constructed libraries was confirmed by 4150 TapeStation System (Agilent) according to the manufacturer’s protocol (High Sensitivity RNA ver.3.0_2021.04). The libraries were sequenced using MiSeq (Illumina) with MiSeq Reagent Kit v3 (600 cycles) (Illumina). TCR clonotypes were quantified with IgBLAST (version 1.20.0) based on the IMGT database (downloaded on 4/12/2023) by Change-O package of Immcantation pipeline (version 1.3.0). Clonal diversity analysis was performed using scikit-bio (version 0.5.6) and other analyses and visualization were performed using matplotlib (version 3.5.1), seaborn (version 0.11.2), ggplot2 (version 3.3.6), and dplyr (version 1.0.9). Python and R versions were 3.9.7 and 4.2.0, respectively.

#### Human leukocyte antigen (HLA) genotyping

RNA extraction was performed using the miRNeasy Micro Kit (Qiagen) according to the manufacturer’s protocol. Centrifugation in the protocol was performed at 8,000 × *g* for “≥8,000 × *g* (≥10,000 rpm)” and at 15,000 rpm for “full speed”. DNase treatment of optional procedure was not performed, but addition of buffer RWT (Step 11) was performed. The extracted RNA was confirmed by 2100 Bioanalyzer Instrument (Agilent) with Agilent RNA 6000 Pico kit (Agilent) according to the manufacturer’s protocol (RNA pico kit manual v01.10). After RNA extraction, cDNA synthesis was performed using SMART-Seq^®^ HT Kit according to the manufacturer’s protocol (Takara Bio Inc.) and the cDNA was amplified for 12 cycles. Library construction was performed using Nextera XT DNA Library Preparation Kit (Illumina) following the manufacturer’s protocol and purified using Agencourt AMPure XP (Beckman Coulter). The libraries were sequenced using Novaseq 6000 (Illumina). In data analysis, sequences were mapped using STAR (version 2.7.2a) with human GRCh38 reference genome after adapter sequences were removed using TrimGalore (version 0.6.6).

HLA genotyping was performed using Optitype (version 1.3.3) and arcasHLA (version 0.5.0) with default parameters.

#### Preparation of TCR cells and APCs

For determination of TCR epitopes and restricting HLA, TCR reconstituted cells and APCs were prepared as described previously.^54,61^ Briefly, TCR reconstituted cells were prepared as reporter cells by introducing TCRα and β chain cDNA sequences into a mouse T-cell hybridoma with a nuclear factor of activated T cells (NFAT)-green fluorescent protein (GFP) reporter gene^62^ using retroviral vectors pMX-IRES-rat CD2. Transformed B cells and HLA-transfected HEK293T cells were generated as APCs. For transformed B cells, 3 × 10^5^ PBMCs were incubated with the recombinant Epstein-Barr virus suspension^63^ for 1 h at 37°C with mild shaking every 15 min. The infected cells were cultured in RPMI 1640 supplemented with 20% FBS (NICHIREI BIOSCIENCES) containing cyclosporine A (CsA, 0.1 μg/mL, Cayman Chemical). Immortalized B lymphoblastoid cell lines were obtained after 3 weeks of culture and used as APCs. For HLA-transfected HEK293T cells, plasmids encoding HLA class I/II alleles^64^ were transfected in HEK293T cells with PEI MAX (Polysciences). For antigen stimulation, TCR-reconstituted cells were co-cultured with 1 μg/mL of peptides in the presence of APCs. After 20 h, cell activation was assessed by GFP and CD69 expression.

#### TCR epitope & HLA restriction

Determination of TCR epitopes and restricting HLA were conducted by using rapid epitope determination platform as described previously.^54,61^ Briefly, 15-mer peptides with 11 amino acids overlap that cover the full length of S protein of SARS-CoV-2 were synthesized (GenScript). Peptides were dissolved in DMSO at 12 mg/mL and 12–15 peptides were mixed to create 26 different semi-pools. TCR-reconstituted reporter cells were stimulated with 1 μg/mL of S peptide pool (1 μg/mL per peptide, JPT), then semi-pools, and then 12 individual peptides in the presence of autologous B cells to identify epitope peptides. To determine the restricting HLA, HLAs were narrowed down by co-culturing reporter cells with autologous and various heterologous B cells in the presence of 1 μg/mL of the epitope peptide. HLAs shared by activating B cells were transduced in HEK239T cells and used for further co-culture to identify the restricting HLA.

#### Outcomes

Primary endpoints included the incidence of AEs/treatment-related AEs/SAEs/solicited AEs, vital signs, laboratory tests, and 12-lead electrocardiograms. Secondary endpoints were related to immunogenicity, including GMT, GMFR, and seroconversion rate (for SARS-CoV-2 NAb titer and anti-spike protein IgG antibodies). Exploratory endpoints included immunological indices such as cytokine-producing cell count to assess cellular immunity, T-cell cytokine assay to assess Th1/Th2 balance, gene analysis of BCRs and TCRs, and biomarker analysis to assess the SARS-CoV-2 NAb titer without using viruses. ELISPOT assay and ICS-flow cytometry assay were performed after cell stimulation by SARS-CoV-2 spike peptide pool.

#### Compliance

The study was conducted in accordance with the protocol (approved by the Institutional Review Board), the Declaration of Helsinki and Council for International Organizations of Medical Sciences (CIOMS) International Ethical Guidelines, the International Council for Harmonization of Technical Requirements for Pharmaceuticals for Human Use (ICH) Good Clinical Practice Guidelines, and other applicable laws and regulations. All participants provided their written informed consent. All authors had access to the study data and reviewed and approved the final manuscript.

### QUANTIFICATION AND STATISTICAL ANALYSIS

The target sample size of up to 60 participants (24 participants × 2 in the S-268019-b groups and 12 participants in the placebo group) was determined based on the precision of safety assessment. The safety assessment precision for each S-268019-b group was calculated based on the probabilities of observing AEs given various true event rates. With 24 participants per group, the probability of observing an AE with a 10% true event rate was 92%, and with a 15% true event rate, it was 98%. Safety analyses were conducted with the safety analysis set which included participants who received at least one dose of the study intervention. AEs were coded and classified by system organ class and preferred term using the Medical Dictionary for Regulatory Activities (MedDRA) Version 24.0.

Immunogenicity was analyzed in the full analysis set which included participants who received at least one dose of the study intervention and who had at least one record of post-vaccination immunogenicity data. Quantitative variables were summarized using mean, SD, median, minimum, and maximum. Categorical variables were summarized using frequency (%). The GMT or GMFR and the corresponding 95% CI were calculated by back transformation of the arithmetic mean and 95% CI of the log-transformed titers or the change from baseline in log-transformed titers, respectively. The 95% CIs for the incidence and the seroconversion rate were calculated using the Clopper-Pearson method. SAS (version 9.4) and R (version 4.0.5) were used for all statistical analyses.

