## Supplementary Figures for "Longitudinal analysis of B- and T-cell responses to SARS-CoV-2 recombinant S-protein vaccine S-268019-b in phase 1/2 priming and booster study"

**SUPPLEMENTARY MATERIAL**

**Figure S1. Immunogenicity of S-268019-b (5 μg and placebo). Related to Figure 2.**

**(A)** and **(B)** GMT for anti-spike protein IgG antibody, in participants receiving 5 μg S‑268019‑b and placebo. **(C)** and **(D)** GMT for SARS‑CoV‑2 neutralizing antibody, in participants receiving 5 μg S‑268019‑b and placebo. **(E)** Percentage of T cells which produces IFN-γ or IL-2 (Th1) cells and IL- 4 or IL-5 (Th2) cells among CD4^+^ T cells by time point.

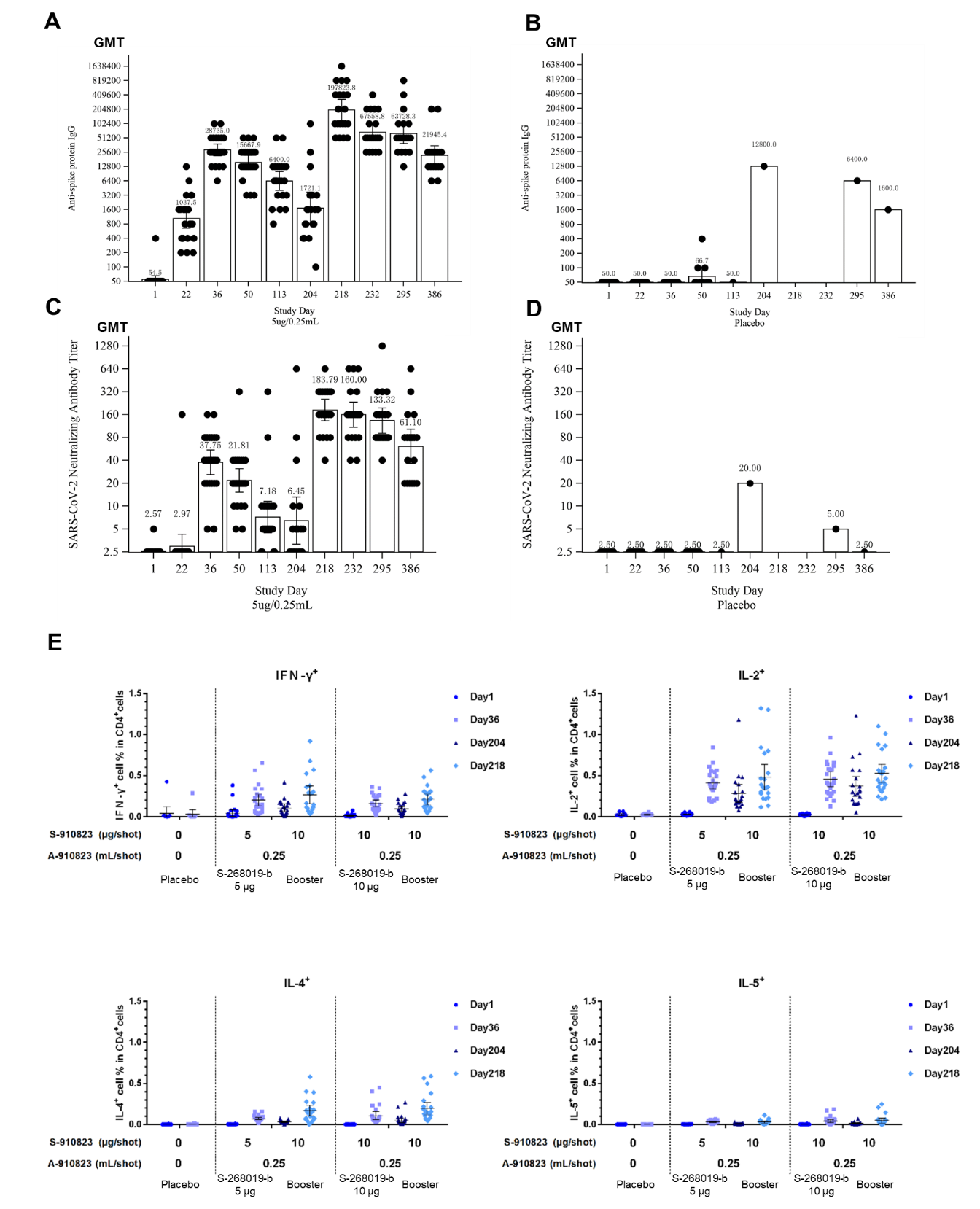

Group = Actual intervention in the primary series. Group names show the antigen content (μg)/adjuvant content (mL).

Abbreviations: GMT, geometric mean titers; IgG, immunoglobulin G; SARS‑CoV‑2, severe acute respiratory syndrome coronavirus 2; IFN-γ, interferon gamma; IL, interleukin.

**Figure S2. Probe validation and gating strategy of B cells. Related to Figure 3.**

**(A)** Confirmation of antigen probe performance to distinguish B cells from other cell types. (**B)** Sorting strategy employed to isolate fraction of B cells. The APC^+^PE^+^ fraction was selected for further analysis.

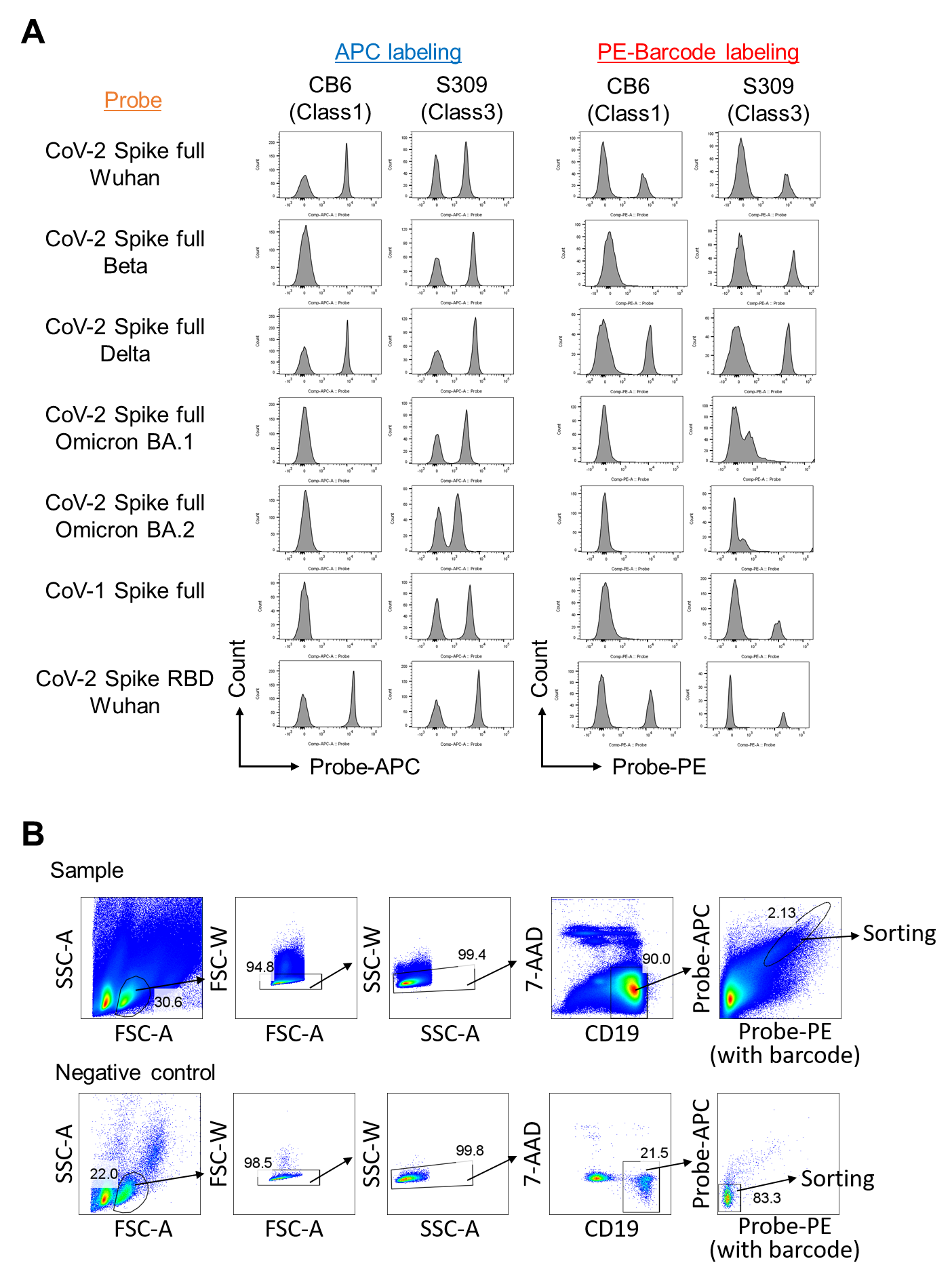

**Figure S3. Single-cell analysis of memory B cells (additional data). Related to Figure 3.**

**(A)** Bar graphs depicting the distribution of different memory B-cell subtypes across various time points. Each bar represents mean ± SD. **(B)** Vector field plots illustrating RNA velocity within single-cell transcriptomics data. **(C)** Isotype distribution (e.g., IgG, IgA, IgM) produced by memory B cells at various time points. **(D)** Graphical representation of the clonal diversity of memory B cells over time. Each point represents the value for each cell. Each bar represents mean ± SD. **(E)** The top panel shows the frequency of different heavy chain variable (V) genes used by memory B cells. The bottom panel provides detailed information on the top 20 most frequently used V genes, including their relative abundances. **(F)** Correlation between Day 50 subtype ratio and neutralizing antibody titer. Statistical analyses were performed using pairwise Wilcoxon signed rank test with multiple comparison correction for (D) and Spearman’s rank-order correlation test for (F) (**p*<0.05, ***p*<0.005, ****p*<0.0005).

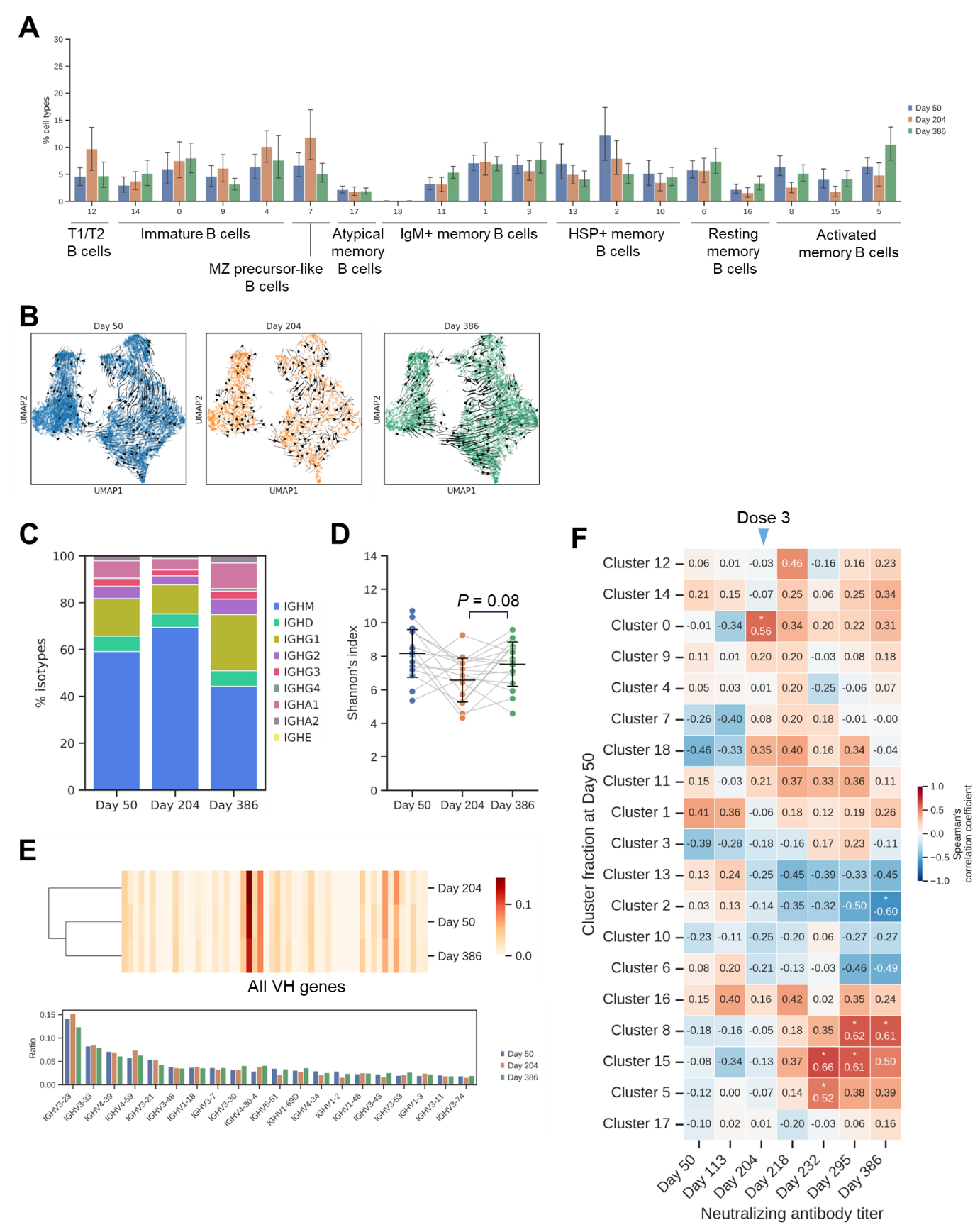

**Figure S4. Epitope mapping of memory B cells (additional data). Related to Figure 4.**

**(A)** A summary of memory B-cell clones that are shared across different time points. **(B)** Representative examples of each class of identified epitopes. **(C)** A detailed summary of the class 3_1 antibodies, including the sequences of the RBD they recognize, the specific epitopes targeted, results from functional assays, and their mapping on the 3D structure of the protein.

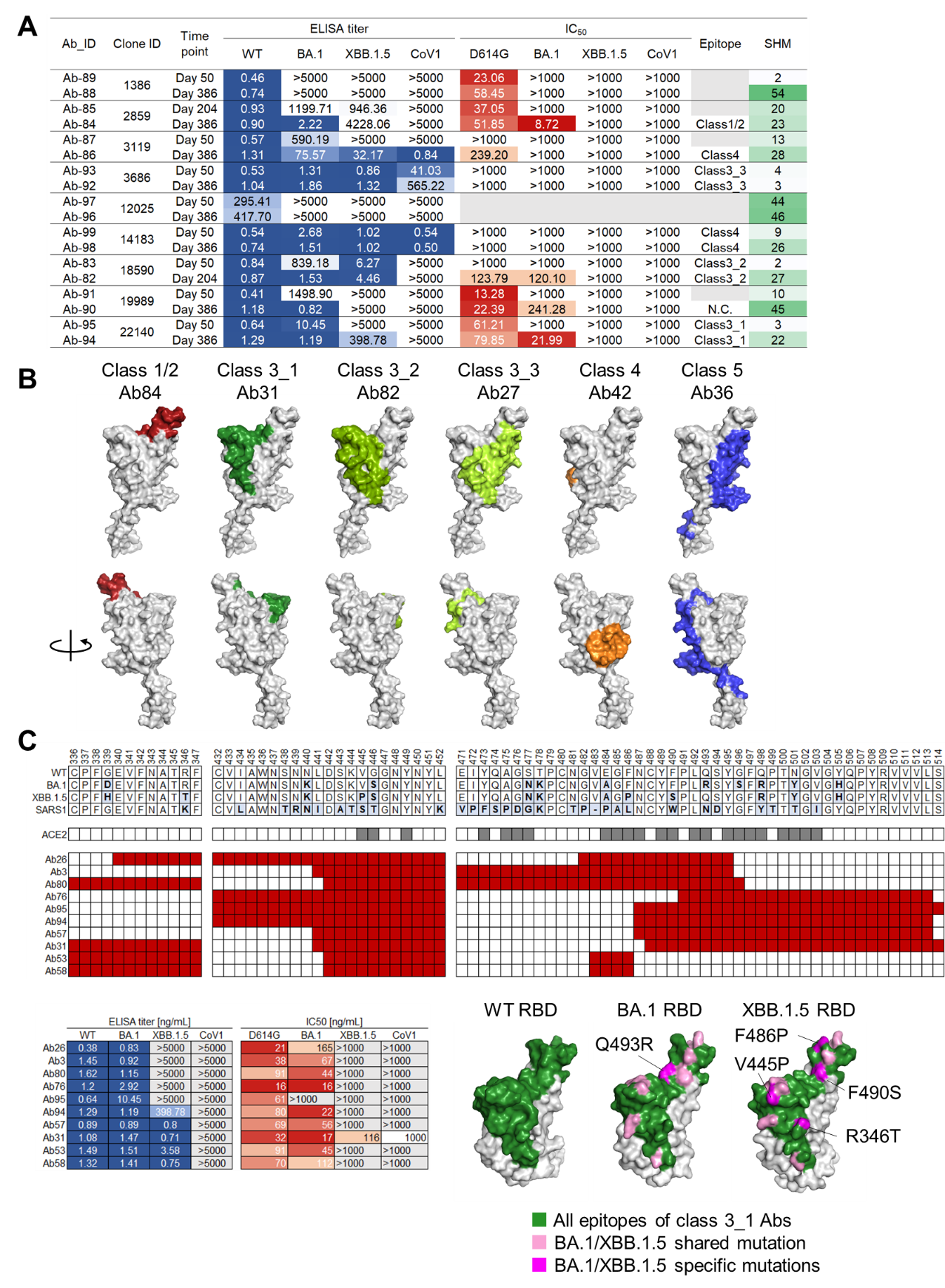

**Figure S5. Single-cell analysis of T cells after priming vaccination (additional data). Related to Figure 5.**

**(A)** Dot plots illustrate the gating strategy employed to isolate this specific fraction of T cells, CD3^+^CTV^−^ fraction used for analysis. **(B)** The fraction of T-cell subtype. Each point represents the value for each donor. The connected lines indicate samples derived from the same donor. Each bar represents mean ± SD. (**C**) Scatter plot showing the percentage of bulk T-cell data that corresponds to clones with identified epitopes. **(D)** Flow diagram detailing the process of aggregating public clones. **(E)** UMAP plot showing the distribution of public clone T cells. **(F)** Chart indicating the distribution of public clone T cells among different donors. **(G)** Information on the epitopes recognized by the public T-cell clones. Statistical analyses were performed using pairwise Wilcoxon signed rank test with multiple comparison correction for (B) (**p*<0.05, ***p*<0.005, ****p*<0.0005).

Abbreviations: Tfh, T follicular helper cells; Teff, effector T cells; Tm, memory T; Treg, T regulatory cells

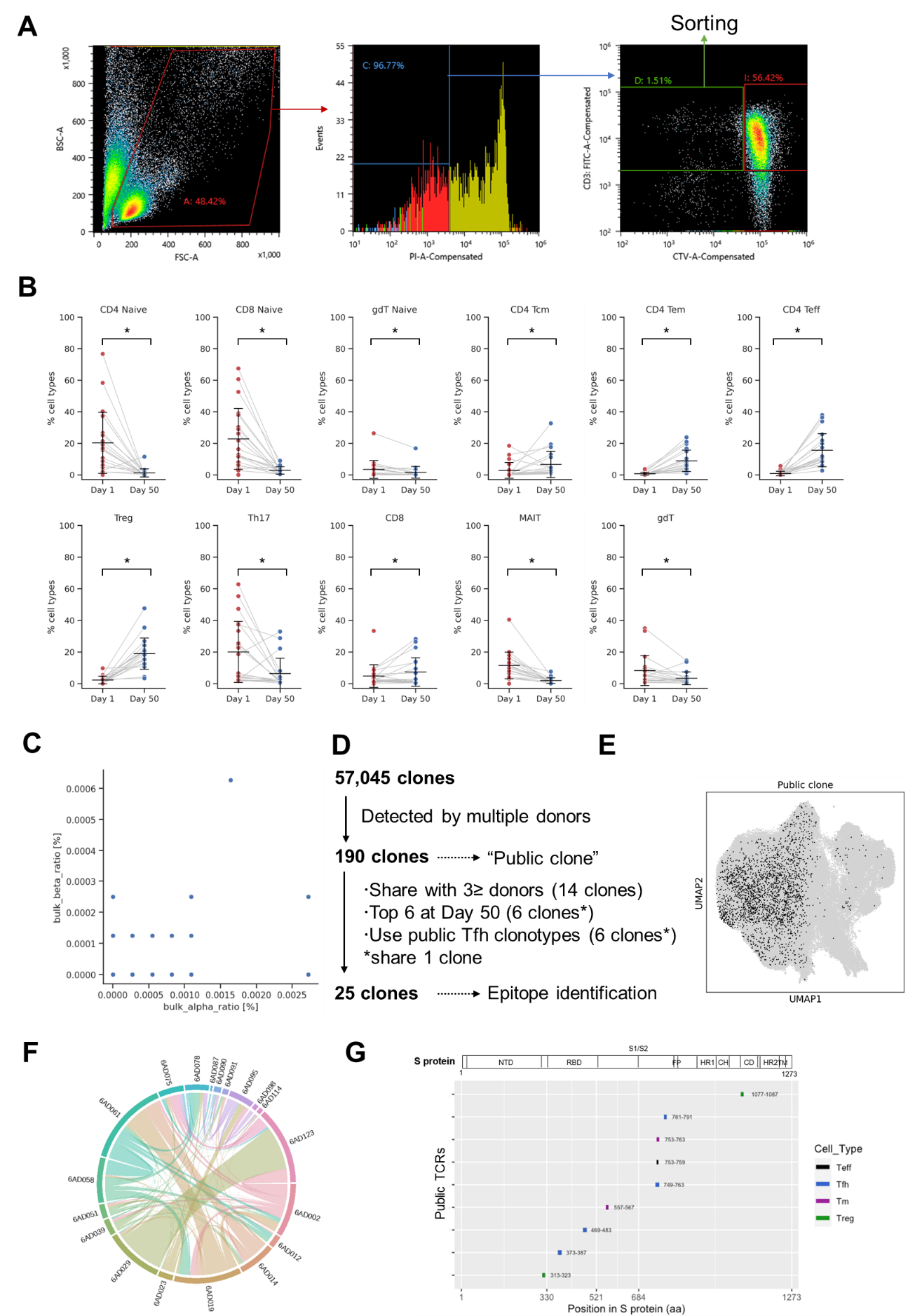

**Figure S6. Single-cell analysis of T cells after booster vaccination (additional data). Related to Figure 6.**

**(A)** Bar graph showing the frequency of different T-cell subtypes at each time point. (B) Bar graphs showing the ratio of different T-cell subtypes at each time point. Each point represents the value for each donor. The connected lines indicate samples derived from the same donor. Each bar represents mean ± SD. **(C)** Bar graphs showing the TCR clonotype, representing the temporal expression patterns of TCRs in response to booster vaccination. **(D)** Graphs presenting the Shannon diversity index for each T-cell subtype. Each bar represents mean ± SD. Statistical analyses were performed using pairwise Wilcoxon signed rank test with multiple comparison correction for (B) (**p*<0.05, ***p*<0.005, ****p*<0.0005).

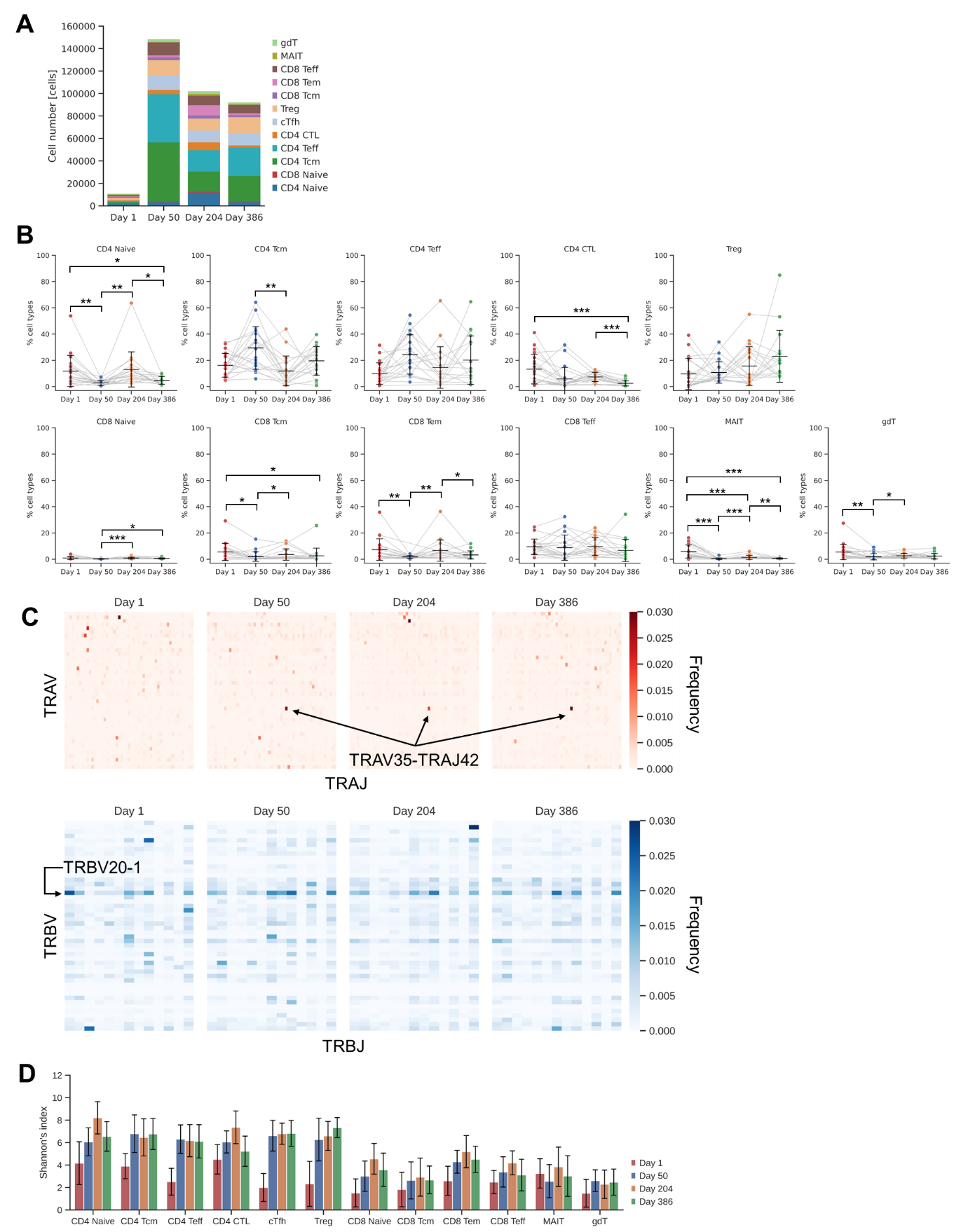

**Table S1. Geometric mean fold rises of anti-spike protein IgG and SARS-CoV-2 neutralizing antibody titers by time point. Related to Figure 2.**

| **Time Point** | **Statistic** | **Anti-spike protein IgG** | | | **SARS-CoV-2 neutralizing antibody titers** | | |
| --- | --- | --- | --- | --- | --- | --- | --- |
|  |  | **5 μg/0.25 mL** | **10 μg/0.25 mL** | **Placebo** | **5 μg/0.25 mL** | **10 μg/0.25 mL** | **Placebo** |
|  |  | ***N* = 24** | ***N* = 24** | ***N* = 12** | ***N* = 24** | ***N* = 24** | ***N* = 12** |
| Day 22 | *n* | 24 | 24 | 12 | 24 | 24 | 12 |
|  | GMFR (95% CI) | 19.03 (12.66–28.59) | 22.63 (14.96–34.23) | 1.00 (–) | 1.16 (0.86–1.56) | 1.03 (0.97–1.09) | 1.00 (–) |
| Day 36 | *n* | 24 | 24 | 12 | 24 | 24 | 12 |
|  | GMFR (95% CI) | 527.00 (375.27–740.10) | 767.13 (555.95–1058.53) | 1.00 (–) | 14.67 (10.25–21.01) | 18.49 (13.89–24.61) | 1.00 (–) |
| Day 50 | *n* | 24 | 24 | 12 | 24 | 24 | 12 |
|  | GMFR (95% CI) | 287.35 (206.47–399.91) | 469.51 (351.21–627.65) | 1.33 (0.90–1.98) | 8.48 (6.14–11.70) | 11.31 (8.61–14.86) | 1.00 (–) |
| Day 113 | *n* | 23 | 22 | 1 | 23 | 22 | 1 |
|  | GMFR (95% CI) | 116.94 (77.82–175.71) | 159.59 (108.71–234.28) | 1.00 (–) | 2.79 (1.81–4.29) | 3.21 (2.44–4.22) | 1.00 (–) |
| Day 204 | *n* | 19 | 22 | 1 | 19 | 22 | 1 |
|  | GMFR (95% CI) | 30.85 (15.19–62.66) | 34.08 (21.43–54.19) | 256.00 (–) | 2.49 (1.25–4.95) | 2.13 (1.40–3.25) | 8.00 (–) |
| Day 218 | *n* | 20 | 22 | 0 | 20 | 22 | 0 |
|  | GMFR (95% CI) | 3565.78 (1937.30–6563.14) | 2896.31 (2092.45–4008.99) | – | 71.01 (51.54–97.85) | 74.92 (58.36–96.17) | – |
| Day 232 | *n* | 20 | 22 | 0 | 20 | 22 | 0 |
|  | GMFR (95% CI) | 1217.75 (736.10–2014.54) | 1237.08 (977.21–1566.06) | – | 61.82 (42.01–90.97) | 66.05 (52.90–82.46) | – |
| Day 295 | *n* | 19 | 22 | 1 | 19 | 22 | 1 |
|  | GMFR (95% CI) | 1142.43 (660.09–1977.24) | 1317.54 (1015.36–1709.66) | 128.00 (–) | 51.42 (34.93–75.69) | 51.33 (41.19–63.97) | 2.00 (–) |
| Day 386 | *n* | 18 | 20 | 1 | 18 | 20 | 1 |
|  | GMFR (95% CI) | 391.02 (224.19–682.02) | 588.13 (388.09–891.28) | 32.00 (–) | 23.52 (14.20–38.95) | 32.00 (21.29–48.10) | 1.00 (–) |

Group = Actual intervention in the primary series. Group names show the antigen content (μg)/adjuvant content (mL).

Abbreviations: CI, confidence interval, GMFR, geometric mean fold rise; IgG, immunoglobulin G; SARS-CoV-2, severe acute respiratory syndrome coronavirus 2.

**Table S3. Definition of RBD epitope class. Related to Figure 4 and STAR Methods.**

| **Epitope class** | **Residue numbers** | **Reference** |
| --- | --- | --- |
| Class 1/2 | 415-421, 452-457, 473-478, 483-494 | [1,2] |
| Class 3 | 333-361, 439-451, 498-501 | [1–5] |
| Class 3_1 | 336-347, 441-452, 487-514 | [1–5] |
| Class 3_2 | 336-361, 441-452 | [1–5] |
| Class 3_3 | 336-361, 453-472 | [1–5] |
| Class 4 | 364-386, 405-408, 412-414, 426-430, 502-504, 515-517 | [2,6] |
| Class 5 | 393-396, 426-430, 462-466, 514-521 | [4] |
| Not classified | 362-363, 387-392, 397-404, 409-411, 422-425, 431-438, 458-461, 467-472, 479-482, 495-497, 505-513, 522-537 |  |
